# Meta-analysis of Genome wide Association Studies on Childhood ADHD Symptoms and Diagnosis Reveals 17 Novel Loci and 22 Potential Effector Genes

**DOI:** 10.1101/2024.04.17.24305817

**Authors:** Camiel M. Van der Laan, Hill F. Ip, Marijn Schipper, Jouke-Jan Hottenga, Eva M. L. Krapohl, Isabell Brikell, María Soler Artigas, Judith Cabana-Domínguez, Llonga Natalia, Ilja M. Nolte, Beate St Pourcain, Koen Bolhuis, Teemu Palviainen, Hadi Zafarmand, Scott Gordon, Tetyana Zayats, Fazil Aliev, Alexandra S. Burt, Carol A. Wang, Gretchen Saunders, Ville Karhunen, Daniel E. Adkins, Richard Border, Roseann E. Peterson, Joseph A. Prinz, Elisabeth Thiering, Natàlia Vilor-Tejedor, Tarunveer S. Ahluwalia, Andrea Allegrini, Kaili Rimfeld, Qi Chen, Yi Lu, Joanna Martin, Rosa Bosch, Josep Antoni Ramos Quiroga, Alexander Neumann, Judith Ensink, Katrina Grasby, José J. Morosoli, Xiaoran Tong, Shelby Marrington, James G. Scott, Andrey A. Shabalin, Robin Corley, Luke M. Evans, Karen Sugden, Silvia Alemany, Lærke Sass, Rebecca Vinding, Erik A. Ehli, Fiona A. Hagenbeek, Eske Derks, Henrik Larsson, Harold Snieder, Charlotte Cecil, Alyce M. Whipp, Tellervo Korhonen, Eero Vuoksimaa, Richard J. Rose, André G. Uitterlinden, Jan Haavik, Jennifer R. Harris, Øyvind Helgeland, Stefan Johansson, Gun Peggy S. Knudsen, Pal Rasmus Njolstad, Qing Lu, Alina Rodriguez, Anjali K. Henders, Abdullah Mamun, Jackob M. Najman, Sandy Brown, Christian Hopfer, Kenneth Krauter, Chandra Reynolds, Andrew Smolen, Michael Stallings, Sally Wadsworth, Tamara Wall, Lindon Eaves, Judy L. Silberg, Allison Miller, Alexandra Havdahl, Sabrina Llop, Maria-Jose Lopez-Espinosa, Klaus Bønnelykke, Jordi Sunyer, Louise Arseneault, Marie Standl, Joachim Heinrich, Joseph Boden, John Pearson, John Horwood, Martin Kennedy, Richie Poulton, Hermine H. Maes, John Hewitt, William E. Copeland, Christel M. Middeldorp, Gail M. Williams, Naomi Wray, Marjo-Riitta Järvelin, Matt McGue, William Iacono, Avshalom Caspi, Terrie E. Moffitt, Andrew Whitehouse, Craig E. Pennell, Kelly L. Klump, Chang Jiang, Danielle M. Dick, Ted Reichborn-Kjennerud, Nicholas G. Martin, Sarah E. Medland, Tanja Vrijkotte, Jaakko Kaprio, Henning Tiemeier, George Davey Smith, Catharina A. Hartman, Albertine J. Oldehinkel, Miquel Casas, Marta Ribasés, Paul Lichtenstein, Sebastian Lundström, Robert Plomin, Meike Bartels, Michel G. Nivard, Dorret I. Boomsma

**Affiliations:** Department of Biological Psychology, Vrije Universiteit Amsterdam, Amsterdam, The Netherlands; Netherlands Institute for the Study of Crime and Law Enforcement; Department of Complex Trait Genetics, Center for Neurogenomics and Cognitive Research, Amsterdam Neuroscience, Vrije Universiteit Amsterdam, Amsterdam, The Netherlands; Institute of Psychiatry, Psychology and Neuroscience, King’s College London, United Kingdom; Department of Medical Epidemiology and Biostatistics, Karolinska Institutet, Stockholm, Sweden; Biomedical Network Research Centre on Mental Health (CIBERSAM), Instituto de Salud Carlos III, Barcelona, Spain; Department of Mental Health, Hospital Universitari Vall d’Hebron, Barcelona, Spain; Psychiatric Genetics Unit, Group of Psychiatry, Mental Health and Addiction, Vall d’Hebron Research Institute (VHIR), Universitat Autònoma de Barcelona, Barcelona, Spain; Department of Epidemiology, University of Groningen, University Medical Center Groningen, Groningen, The Netherlands; MRC Integrative Epidemiology Unit, University of Bristol, Bristol, UK; Max Planck Institute for Psycholinguistics, The Netherlands; Donders Institute for Brain, Cognition and Behaviour, Radboud University, The Netherlands; Department of Child and Adolescent Psychiatry/Psychology, Erasmus University Medical Center, Rotterdam, Netherlands; Institute for Molecular Medicine FIMM, HiLife, University of Helsinki, Helsinki, Finland; Department of Public Health, Amsterdam Public Health Research Institute, Amsterdam UMC, location Academic Medical Center, University of Amsterdam, Amsterdam, The Netherlands; Department of Clinical Epidemiology, Biostatistics and Bioinformatics, Amsterdam Public Health Research Institute, Amsterdam UMC, location Academic Medical Center, University of Amsterdam, Amsterdam, The Netherlands; QIMR Berghofer Medical Research Institute, Brisbane, Australia; K.G. Jebsen Centre for Neuropsychiatric Disorders, Department of Biomedicine, University of Bergen, Norway; Analytic and Translational Genetics Unit, Department of Medicine, Massachusetts General Hospital and Harvard Medical School, Boston, Massachusetts, USA; Stanley Center for Psychiatric Research, Broad Institute of MIT and Harvard, Cambridge, Massachusetts, USA; Department of Psychiatry, Robert Wood Johnson Medical School, Rutgers University, U.S.A; Department of Psychology, Michigan State University, East Lansing, USA; School of Medicine and Public Health. Faculty of Medicine and Health, The University of Newcastle, Newcastle, New South Wales, Australia; Hunter Medical Research Institute, Newcastle, New South Wales, Australia; Department of Psychology, University of Minnesota, USA; MRC Centre for Environment and Health, Department of Epidemiology and Biostatistics, School of Public Health, Imperial College London, London, United Kingdom; Faculty of Medicine, University of Oulu, PO Box 8000, FI-90014 Oulun yliopisto, Finland; Department of Sociology, College of Social and Behavioral Science, University of Utah; Department of Psychiatry, School of Medicine, University of Utah; Departments of Neurology and Computer Science, University of California Los Angeles, California, USA; Department of Psychiatry and Behavioral Sciences, State University of New York Downstate Health Sciences University, Brooklyn, NY, USA; Department of Psychiatry, Virginia Institute for Psychiatric and Behavioral Genetics, Virginia Commonwealth University, Richmond, VA, USA; Center for Genomic and Computational Biology, Duke University, Durham, NC, USA; Institute of Epidemiology, Helmholtz Zentrum München - German Research Center for Environmental Health, Neuherberg, Germany; Ludwig-Maximilians-University of Munich, Dr. von Hauner Children’s Hospital, Division of Metabolic Diseases and Nutritional Medicine, Munich, Germany; BarcelonaBeta Brain Research Center, Pasqual Maragall Foundation (FPM), Barcelona, Spain; Centre for Genomic Regulation (CRG), The Barcelona Institute of Science and Technology, Barcelona, Spain; IMIM (Hospital del Mar Medical Research Institute), Barcelona, Spain; Department of Clinical Genetics. Erasmus University Medical Center. Rotterdam, Netherlands; COPSAC, Copenhagen Prospective Studies on Asthma in Childhood, Herlev and Gentofte Hospital, University of Copenhagen, Copenhagen, Denmark; Steno Diabetes Center Copenhagen, Herlev, Denmark; SJD MIND Schools Program, Hospital Sant Joan de Déu, Institut de Recerca Sant Joan de Déu, Esplugues de Llobregat, Spain; Department of Psychiatry and Legal Medicine, Universitat Autònoma de Barcelona, Barcelona, Spain; Department of child and adolescent psychiatry and psychology, Amsterdam UMC, Amsterdam; Reproduction and Development research Institute, Amsterdam, the Netherlands; Amsterdam Public Health Research Institute, Amsterdam, the Netherlands; Levvel, Academic center for Child and Adolescent Psychiatry, Amsterdam, The Netherlands; Department of Clinical, Educational, and Health Psychology, University College London, London, UK; Mental Health and Neuroscience Research Program, QIMR Berghofer Medical Research Institute, Brisbane, Australia; School of Psychology, University of Queensland, Brisbane, Australia; Center for Innovation in Public Health, University of Kentucky; School of Public Health, The University of Queensland, Herston, Qld, Australia; Child Health Research Centre, The University of Queensland, Brisbane, QLD, Australia; Child and Youth Mental Health Service, Children’s Health Queensland Hospital and Health Service, Brisbane, QLD, Australia; Queensland Centre for Mental Health Research, QLD, Australia; Institute for Behavioral Genetics, University of Colorado Boulder, Colorado, USA; Department of Ecology and Evolutionary Biology, University of Colorado Boulder, Colorado, USA; Department of Psychology and Neuroscience, Duke University, Durham, N.C., U.S.A; Avera Institute for Human Genetics, Sioux Falls, South Dakota, USA; Translational Neurogenomics Laboratory, QIMR Berghofer Medical Research Institute, Brisbane, Queensland, Australia; School of Medical Sciences, Orebro University, Orebro, Sweden; Department of Biomedical Data Sciences, Molecular Epidemiology, Leiden University Medical Center, Leiden, The Netherlands; Department of Epidemiology, Erasmus University Medical Center, Rotterdam, The Netherlands; Department of Psychological and Brain Sciences, Indiana University, Bloomington, IN, USA; Department of Internal Medicine, Erasmus University Medical Center, Rotterdam, The Netherlands; Netherlands Genomics Initiative (NGI)-sponsored Netherlands Consortium for Healthy Aging (NCHA), Leiden, The Netherlands; Centre for Fertility and Health, The Norwegian Institute of Public Health, Oslo, Norway; Department of Genetics and Bioinformatics, Division of Health Data and Digitalization, The Norwegian Institute of Public Health, Bergen, Norway; Department of Clinical Science, University of Bergen, Norway; Department of Biostatistics, University of Florida, Gainesville, USA; Institute for Molecular Bioscience, University of Queensland, QLD, Australia; Institute for Social Science Research, University of Queensland, QLD, Australia; Department of Psychiatry, University of California, San Diego; University of Colorado School of Medicine, Aurora, Colorado, USA; Department of Molecular, Cellular, and Developmental Biology, University of Colorado Boulder, USA; Department of Psychology, University of California Riverside, Riverside, CA, USA; Department of Psychology and Neuroscience, University of Colorado Boulder, Colorado, USA; Department of Human and Molecular Genetics, Virginia Commonwealth University, Richmond, VA, USA; Department of Pathology and Biomedical Science, and Carney Centre for Pharmacogenomics, University of Otago Christchurch, New Zealand; PsychGen Centre for Genetic Epidemiology and Mental Health, Norwegian Institute of Public Health, Oslo, Norway; Nic Waals Institute, Lovisenberg Diaconal Hospital, Oslo, Norway; Epidemiology and Environmental Health Joint Research Unit, FISABIO-Universitat Jaume I- Universitat de València, Valencia (Spain); Spanish Consortium for Research on Epidemiology and Public Health (CIBERESP), Madrid (Spain); Department of Nursing, Universitat de València, Valencia, Spain; ISGlobal, Barcelona Institute for Global Health, Barcelona, Spain; Universitat Pompeu Fabra (UPF), Barcelona, Spain; CIBER Epidemiología y Salud Pública (CIBERESP), Spain; Social, Genetic and Developmental Psychiatry Centre, Institute of Psychiatry, Psychology and Neuroscience, King’s College London, U.K; Institute and Outpatient Clinic for Occupational, Social and Environmental Medicine, University of Munich Medical Center, Ludwig-Maximilians-Universität München, Munich, Germany; Allergy and Lung Health Unit, Melbourne School of Population and Global Health, University of Melbourne, Melbourne, Australia; Christchurch Health and Development Study, Department of Psychological Medicine, University of Otago Christchurch, New Zealand; Biostatistics and Computational Biology Unit, Department of Pathology and Biomedical Science, University of Otago Christchurch, New Zealand; Dunedin Multidisciplinary Health and Development Research Unit, University of Otago, Dunedin, New Zealand; Massey Cancer Center, Virginia Commonwealth University, Richmond, VA, USA; Department of Psychiatry, College of Medicine, University of Vermont; Arkin mental health care, Amsterdam, The Netherlands; Queensland Brain Institute, Institute for Molecular Bioscience, University of Queensland, St Lucia 4072, Australia; Department of Life Sciences, College of Health and Life Sciences, Brunel University London, London, United Kingdom; Department of Psychiatry and Behavioral Sciences, Duke University School of Medicine, Durham, N.C., U.S.A; Center for Genomic and Computational Biology, Duke University, Durham, N.C., U.S.A; Telethon Kids Institute, University of Western Australia, Nedlands, Western Australia, Australia; Department of Psychology, Virginia Commonwealth University, U.S.A; College Behavioral and Emotional Health Institute, Virginia Commonwealth University, Richmond, VA, USA; Institute of Clinical Medicine, University of Oslo, Oslo, Norway; School of Psychology and Counselling, Queensland University of Technology, Brisbane, Australia; Department of Public and Occupational Health, Amsterdam University Medical Center, University of Amsterdam, Amsterdam; Amsterdam Reproduction and Development Research Institute, Amsterdam, The Netherlands; Department of Public Health, Medical Faculty, University of Helsinki, Helsinki, Finland; Department of Social and Behavioral Science, Harvard TH Chan School of Public Health, Boston, USA; Population Health Sciences, Bristol Medical School, University of Bristol, Bristol, UK; Interdisciplinary Center Psychopathology and Emotion regulation (ICPE), Department of Psychiatry, University of Groningen, University Medical Center Groningen, Groningen, The Netherlands; Fundació d’Investigació Sant Pau, Barcelona, Spain; Instituto para el Desarrollo de Terapias Avanzadas en Neurociencias (IDETAN). Barcelona, Spain; Gillberg Neuropsychiatry Centre, University of Gothenburg, Gothenburg, Sweden; Centre for Ethics, Law and Mental Health, University of Gothenburg, Gothenburg, Sweden

**Keywords:** ADHD, GWAS, symptoms, diagnosis, genetics, FLAMES

## Abstract

Attention-deficit/hyperactivity disorder (ADHD) is a heritable neurodevelopmental disorder for which genetic factors explain up to 75% of the variance. In this study, we performed a genome-wide association meta-analysis (GWAMA) of ADHD symptom measures, with an effective sample size of 120,092 (71,733 unique individuals from 28 population-based cohorts, with 288,887 quantitative ADHD symptom measures). Next, we meta-analyzed the results with a genome-wide association study (GWAS) of ADHD diagnosis. The GWAMA of ADHD symptoms returned no genome-wide significant variants. However, we estimated strong genetic correlations between our study of quantitative ADHD symptoms and the earlier study of ADHD diagnosis (*r*_g_= 1.00, SE= 0.06). Moderate negative genetic correlations (*r*_g_< -0.40) were observed with several cognitive traits. Genetic correlations between ADHD and aggressive behavior and antisocial behavior were around 1. This provides further evidence of the wide pleiotropic effects of genetic variants and the role that genetic variants play in the co-occurrence with (mental) health traits. The GWAMAs of ADHD symptoms and diagnosis identified 2,039 genome-wide significant variants, representing 39 independent loci, of which 17 were new. Using a novel fine-mapping and functional annotation method, we identified 22 potential effector genes which implicate several new potential biological processes and pathways that may play a role in ADHD. Our findings support the notion that clinical ADHD is at the extreme end of a continuous liability that is indexed by ADHD symptoms. We show that including ADHD symptom counts in large-scale GWAS can be useful to identify novel genes implicated in ADHD and related symptoms.

## Introduction

Attention-deficit/hyperactivity disorder (ADHD) is, for many individuals, a persistent neurodevelopmental disorder (Faraone et al., 2006; Kan et al., 2013). ADHD is characterized by three core symptoms: hyperactivity, impulsivity, and inattention (American Psychiatric Association, 2013). It affects around 5% of children and adolescents and 2.5% of adults worldwide (Faraone et al., 2015). ADHD may be associated with serious consequences for affected individuals, their families, and society at large, with symptoms persisting across multiple settings, i.e. at home, at school, and elsewhere (Caci et al., 2014, 2015). The disorder has a predominantly genetic aetiology, involving both common and rare genetic variants (Faraone et al., 2024). The mean estimated heritability across 37 twin studies of ADHD was 74% (Faraone & Larsson, 2019; Kan et al., 2014; Merwood et al., 2013), with some differences in heritability across ages and raters (Kan et al., 2014; Merwood et al., 2013).

In 2019, a genome-wide association meta-analysis (GWAMA) of clinical ADHD, hereafter referred to as ADHD_DIAG2019_, including data from 20,183 cases and 35,191 controls, identified the first 12 genome-wide significant loci associated with ADHD (Demontis et al., 2019). The study reported that 22% of the variance in ADHD could be explained by all measured single nucleotide polymorphisms (SNPs). They also performed meta-analyses with deCode, 23andMe, and EAGLE (Middeldorp et al., 2016). Four independent loci reached the genome-wide significance threshold (*p*< 5×10^-8^) in all three meta-analyses. Interestingly, most independent significant loci, 15, were found in the meta-analysis with a quantitative assessment of attention problems in EAGLE, suggesting that quantitative assessments of ADHD can boost the power to identify implicated genetic variants. Ten independent significant loci were found in meta-analyses with deCode and 23andMe data. In 2023, Demontis and colleagues presented results from their updated GWAS meta-analysis of ADHD (ADHD_DIAG_), combining newly extended data from iPSYCH, deCode, and the PGC, almost doubling the number of cases compared with ADHD_DIAG2019_ (Demontis et al., 2023). The definition of cases was broader, for example by including individuals who used ADHD prescription medication. The study reported 27 independent significant loci and estimated that 14% of the variance in ADHD could be attributed to the included SNPs. The broader definition of ADHD diagnosis resulted in a larger sample and, therefore, more power to detect implicated genetic variants. However, this broader definition also increased the heterogeneity of the phenotype, which may explain the decrease in SNP-heritability (Wang et al., 2023).

There is increasing interest and recognition that ADHD symptom counts in non-clinical samples can tap into the same genetic construct as clinically diagnosed ADHD, supporting the notion that ‘clinical ADHD’ is at the extreme end of a continuous measure of ADHD symptoms (Larsson et al., 2012; Levy et al., 1997). This was initially suggested based on multivariate twin studies (Derks et al., 2008). Additionally, the genetic correlation (*r*_g_) between quantitative ADHD symptom counts (Middeldorp et al., 2016) and ADHD_DIAG2019_ (Demontis et al., 2019) was estimated to be 0.97 (SE= 0.21, *p*= 2.66 x 10^-6^), suggesting that combining these measures is a viable strategy to increase statistical power in ADHD GWASs. This was additionally supported by the increased number of genome-wide significant loci in the meta-analysis of ADHD_DIAG2019_ and EAGLE, as compared to ADHD_DIAG2019_ alone, and to meta-analyses of ADHD_DIAG2019_, deCode and 23andMe.

In this study, we combine information from 28 population-based cohorts in a GWAMA of continuous ADHD symptom scores, leading to a total of 71,733 participants. The measures include repeated assessments (longitudinal data) by multiple raters (maternal, paternal, teachers, and self-assessments) and instruments across ages (range: 2-18 years), for a total of 288,887 measures. We also include retrospective self-report data. We meta-analyze all available data into a cross-rater/cross-age/cross-instrument GWAMA of ADHD symptoms (ADHD_symp_), taking into consideration the dependency between multiple assessments within individuals (Ip et al., 2021). Using measures from multiple raters and ages may further increase the power of the analyses due to an increase in the validity of the ADHD symptom measures. Next, we estimate the genetic correlations (*r*_g_) between ADHD_SYMP_ and the meta-analysis of case-control samples (Demontis et al., 2023), and meta-analyze ADHD_SYMP_ with ADHD_DIAG_ (ADHD_overall_). Finally, we performed fine mapping and gene-based tests based on ADHD_SYMP_ and ADHD_overall_, performed several follow-up enrichment and pathway analyses, and estimated genetic correlations between the GWAMA and a set of predefined outcomes from cognitive and externalizing behaviour domains.

## Results

### ADHD_SYMP_ GWAMA

We first meta-analyzed the effect of each SNP across all available univariate GWAS of quantitative ADHD measures. Based on an effective sample size of 120,092, the estimated 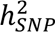 of ADHD_SYMP_ was 0.04 (SE= 0.01; Z= 8.12). The mean *χ*^2^ statistic was 1.09 along with an LDSC-intercept of 1.01 (SE= 0.01), indicating that there was no or very limited inflation in test statistics due to confounding biases such as population stratification. Rather, the GWAMA most likely captured the polygenic nature of childhood ADHD symptoms. The GWAMA of ADHD symptoms did not result in any genome-wide significant SNPs (Figure 1, Supplements Table 11).

**Figure 1.**
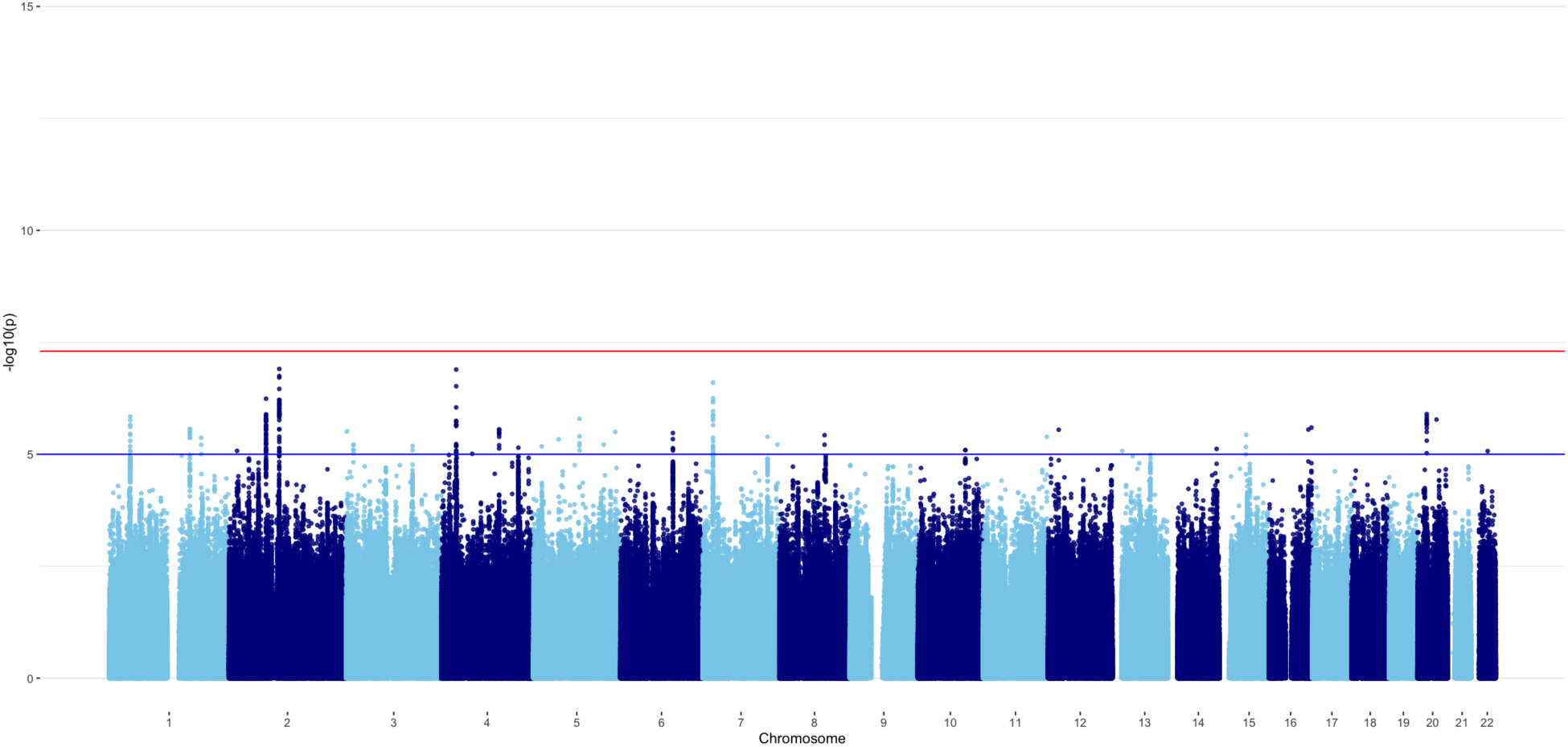
Manhattan plot: GWAMA of ADHD symptoms.

### Meta-analysis with ADHD diagnosis GWAS

SNP-heritability estimated with genomicSEM was 0.13 (SE= 0.01) for ADHD_DIAG_. The estimated genetic correlation between ADHD_SYMP_ and ADHD_DIAG_ was 1.00 (SE= 0.06). The CTI was not significantly different from zero and were subsequently constrained to zero in the following meta-analysis.

Because the point estimate of the genetic correlation between ADHD_SYMP_ and ADHD_DIAG_ was not significantly different from 1, we constrained the genetic correlation at unity when pre-adjusting the weights and Z-scores for the meta-analysis of ADHD_SYMP_ and ADHD_DIAG_. A total of 6,571,852 SNPs were included in the meta-analysis. The SNP-heritability of ADHD_overall_ was 0.11 (SE= 0.01), with a mean *χ*^2^ statistic of 1.52. The LDSC-intercept and ratio were 1.02 (SE= 0.01) and 0.03 (SE= 0.02), respectively, indicating that approximately 3% of the signal might be due to confounding factors. A Manhattan plot of ADHD_overall_ is shown in Figure 2. 2,039 SNPs reached genome-wide significance (*p*< 5 x 10^-8^), of which 644 were also reported in ADHD_DIAG_ and 1,395 were new. The 2,039 SNPs corresponded to 43 independent lead SNPs in 39 independent significant loci, identified with FUMA. Of these 39 loci, 22 were also reported in ADHD_DIAG_ and 17 were new. See also Figure 2 and Supplements Tables 12 and 13.

**Figure 2.**
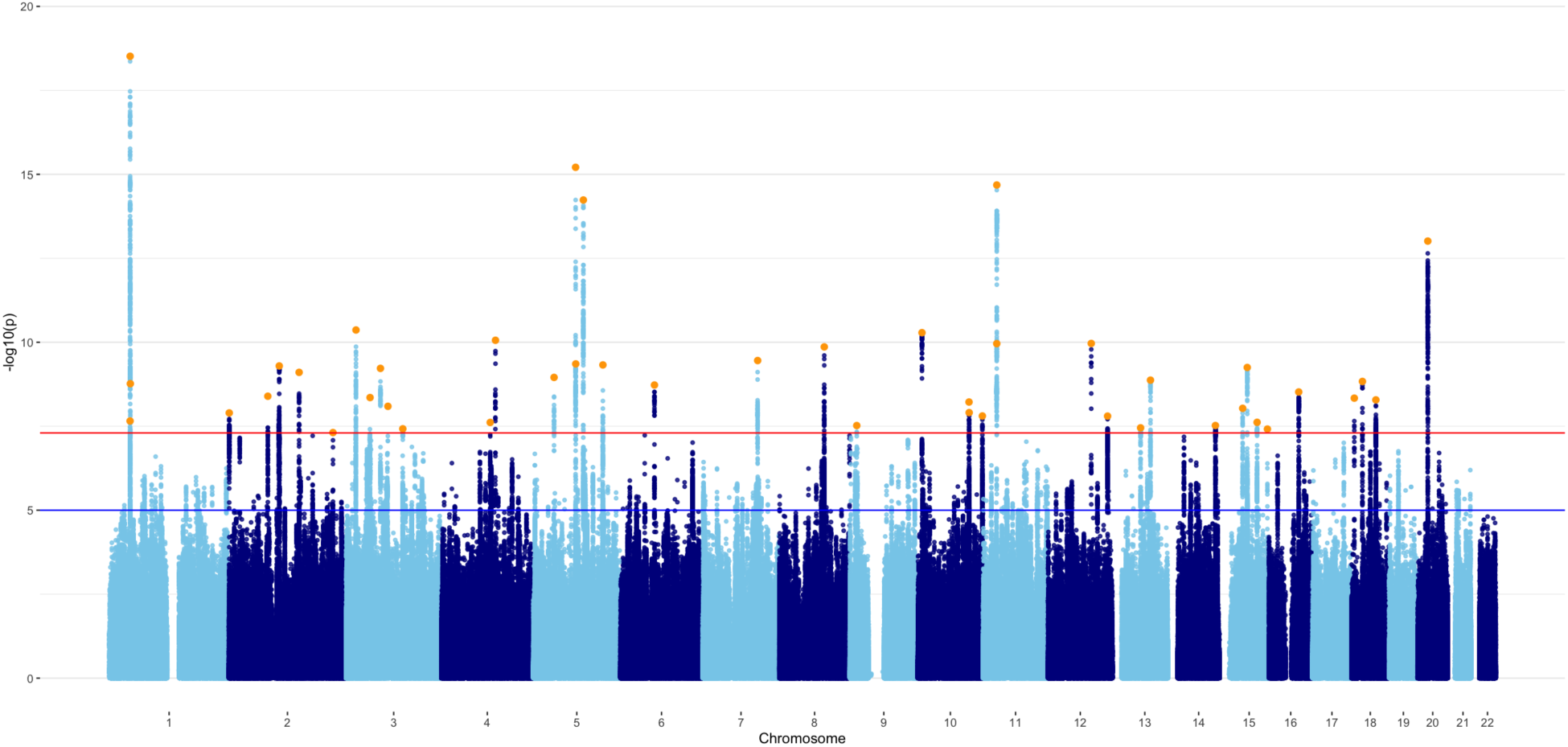
Manhattan plot: GWAMA of ADHD symptoms and ADHD diagnosis. *Note: orange dots reflect lead SNPs*.

### Follow-up analyses

#### Fine mapping & gene-based tests

Gene mapping in FUMA mapped the 43 lead SNPS in 39 independent genomic risk loci to 204 associated genes (See Supplements Table 14), of which 45 were also reported by Demontis et al (2023). Second, gene-based tests were run in MAGMA (de Leeuw et al., 2015), identifying 64 associated genes (see Supplements Table 15), of which 17 were previously reported by Demontis et al. (2023). Third, we ran FLAMES (Schipper et al., 2023), with the aim to get a better understanding of the genes that are causally involved in ADHD. A total of 22 genes had FLAMES scores larger than 0.05, and were interpreted as potential effector genes (see Supplements Table 16). Fourteen of these effector genes were also tagged by the MAGMA gene-based test. Ten of these potential effector genes were previously reported by Demontis et al. (2023). Four genes were not reported by Demontis et al. (2023) but have previously been linked to ADHD, as listed by GWAS Catalog. Eight potential effector genes were not reported by Demontis et al. (2023) or by any ADHD specific studies listed on GWAS Catalog: *EMCN*, *STK32C*, *PCDH17*, *TCF12*, *PEAK1*, *IGF1R*, *CTNNA2*, and *ABCA12*.

#### Enrichment & tissue specific expression

Gene-set analysis in MAGMA revealed no significant enrichment in any MSigDB v2023 gene-sets after correction for multiple testing. MAGMA expression analysis showed significant enrichment of the GWAMA signal in gene-sets differentially expressed in late infancy. Additionally, there was significant enrichment in several brain tissue types, as well as in the pituitary gland (See Supplements Figures 1 and 2).

Next, FUMA GENE2FUNC gene-set enrichment analyses of the 204 potential ADHD risk genes mapped by FUMA showed significant enrichment in genes identified in GWAS of ADHD, cognition-related phenotypes, and risk-taking behaviors. The genes were not significantly enriched in any tissue types, or in any of the Brainspan developmental stages of brain samples. Genes were enriched in 29 gene sets that code for transcription factor targets. Zero synpase cellular component terms or biological processes were significantly enriched at 1% FDR (testing terms with at least three matching input genes in SynGO). For a complete overview of all enrichment results and the included gene sets, see Supplemental Figures 3-5, and Supplemental Notes.

We repeated the same analyses for the 22 potential effector genes identified by FLAMES. Again, genes were highly enriched for genes identified in GWAS of cognition-related phenotypes, and risk-taking behaviors. The 22 genes were significantly overrepresented in gene sets that are differentially expressed in the frontal cortex, but not in the “general tissue” type brain, or in any of the Brainspan developmental stages. Genes were also enriched in 52 gene sets that code for transcription factor targets, 13 microRNA targets, 4 gene ontology biological processes, 1 canonical pathway, and 8 cell type signatures. Zero synpase cellular component terms or biological processes were significantly enriched at 1% FDR (testing terms with at least three matching input genes in SynGO). Nine genes were mapped to SynGO annotations, eight to cellular components and 9 to biological processes. Genes were enriched in integral components of the postsynaptic density membrane (*q*= 1.46 × 10^−3^), postsynaptic density (*q*= 1.57 × 10^−3^), postsynapse (*q*= 5.67 × 10^−3^), and synapse (*q*= 7.22 × 10^−3^), as well as in postsynaptic modulation of chemical synaptic transmission (*q*= 3.28 × 10^−5^), process in the synapse (*q*= 3.97 × 10^−4^), and synapse organization (*q*= 1.62 × 10^−3^). For a complete overview of all enrichment results and the included gene sets, see Supplemental Figures 6-12, Supplementary Tables 17 and 18, and Supplemental Notes.

#### Genetic correlations

We estimated genetic correlations between ADHD_overall_ and 49 external phenotypes. Results are shown in Figure 4 and Supplementary Table 18. Strong positive genetic correlations were observed between ADHD_overall_ and childhood aggressive behavior (*r*_g_= 1.13, SE= 0.05) and antisocial behavior (*r*_g_= 0.97, SE= 0.06). The correlation of 1.13 with childhood aggressive behavior reflects a high genetic correlation that is likely greater than 1 due to sampling variation, correlations estimated by LD score regression in genomicSEM are not bounded between -1 and 1 (Childhood aggression *h^2^ Z*= 9.03). Measures of smoking habits (*r*_g_= 0.46-0.6, SE= 0.03) and number of children (*r*_g_= 0.38, SE= 0.04) also showed moderate correlations, as did ratings of overall health (*r*_g_= -0.59, SE= 0.03), educational attainment (*r*_g_= -0.55, SE= 0.02), and childhood IQ (*r*_g_= -0.43, SE= 0.06). In general, ADHD_overall_ showed weak to moderate genetic correlations with psychopathologies, including Major Depressive Disorder (*r*_g_= 0.57, SE= 0.03) and Autism Spectrum Disorder (*r*_g_= 0.39, SE= 0.04). Weak negative genetic correlations were found between ADHD_overall_ and alcohol intake frequency (*r*_g_= -0.28, SE= 0.03), but a weak positive correlation with drinks per week (*r*_g_= 0.14, SE= 0.03) and a weak positive correlation with cannabis use (*r*_g_= 0.2, SE= 0.03). Marees and colleagues (Marees et al., 2020) looked into similar genetic correlations for alcohol use, and found evidence to suggest that they are the result of SES influences. ADHD_overall_ was weakly negatively genetically correlated with birth weight (*r*_g_= -0.1, SE= 0.02), but showed a weak positive genetic correlation with childhood obesity (*r*_g_= 0.21, SE=0.05) and a moderate positive genetic correlation with adult BMI (*r*_g_= 0.3, SE= 0.02).

**Figure 4.**
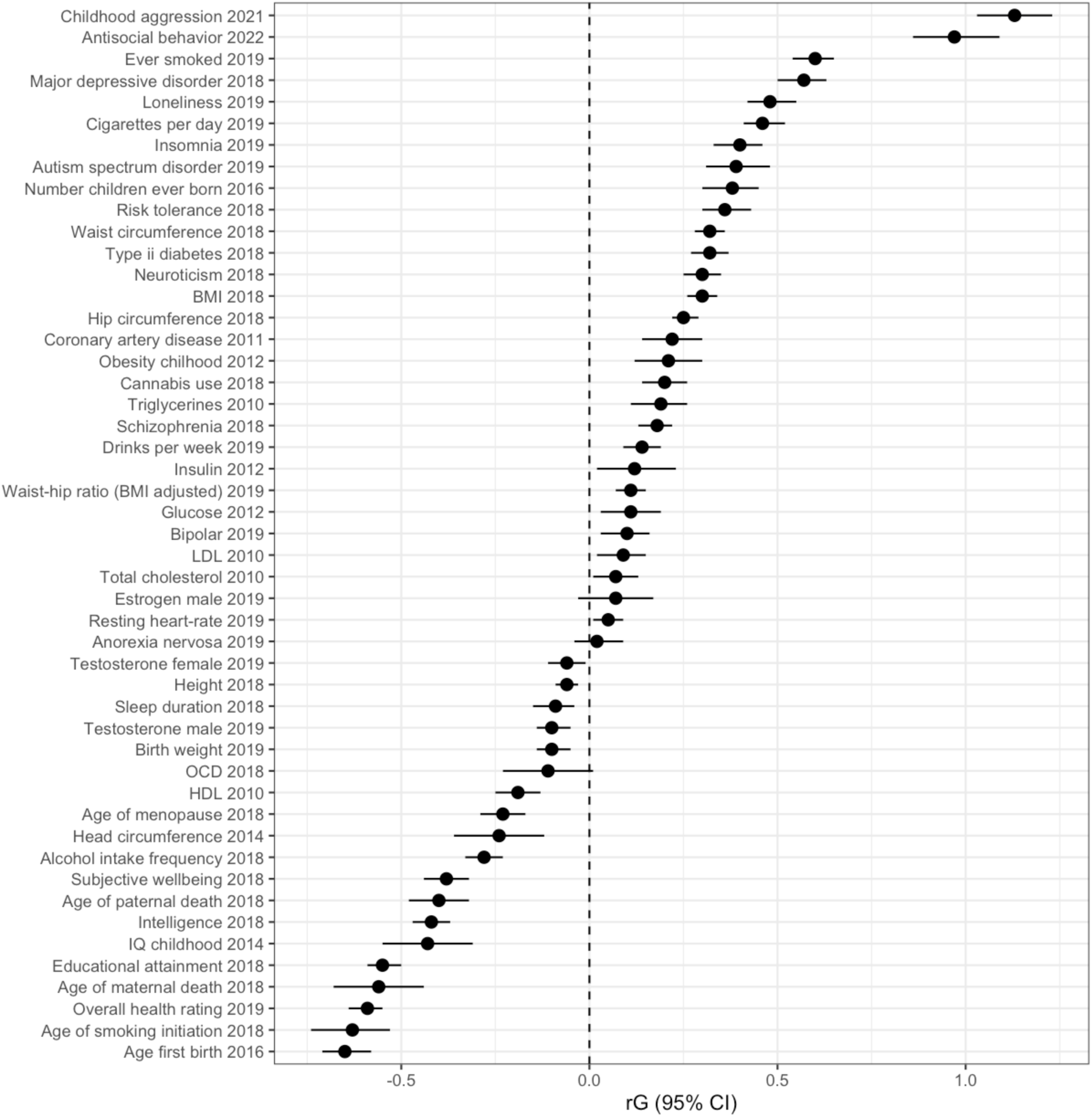
Genetic correlations with external phenotypes. Bars indicate 95% confidence intervals.

## Discussion

We present a genome-wide association meta-analysis of childhood Attention-Deficit/Hyperactivity Disorder symptoms (ADHD_SYMP_). A total of 28 cohorts with measures of ADHD symptom counts took part; contributing data from multiple raters and instruments, across a wide range of ages. We meta-analyzed all continuous measures and combined these results with results from two GWAMAs of ADHD diagnosis (ADHD_DIAG_) by Demontis and colleagues (2019, 2023).

We did not identify any genome-wide significant hits for ADHD symptoms specifically. Genetic correlations with ADHD diagnosis was *r*_g_= 1.00, SE= 0.06. This supports the notion that clinical ADHD is at the extreme end of a continuous genetic liability that is indexed by ADHD symptoms (Larsson et al., 2012; Levy et al., 1997). This was previously suggested based on multivariate twin studies (Derks et al., 2008). The estimated 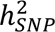 of ADHD_SYMP_ was 0.04 (SE= 0.01), which may be considered low compared to the estimated 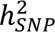 in Demontis et al., 2019 and 2023: 0.22 and 0.14 respectively. This is probably because we introduced heterogeneous measurement error and bias in our phenotyping by including symptom measures from different raters, and at different ages, which could subsequently suppress SNP-heritability.

By meta-analyzing GWASs of ADHD symptoms and ADHD diagnosis, we found 2,039 genome-wide significant variants in 39 independent loci, of which 17 were new. Demontis et al. found 12 and 27 independent hits in 2019 and 2023, which included data from 23andMe. These 23andMe data were excluded from the current meta-analyses. This shows that, because ADHD symptom measures have been widely collected, combining ADHD symptom counts and diagnosis can be effective in identifying implicated genetic variants for ADHD. The estimated 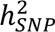 of ADHD_overall_ was 0.11 (SE= 0.01), compared to 0.13 (SE= 0.01) in ADHD_DIAG_ when excluding 23andMe data. This means that by including ADHD_SYMP_ in the ADHD_DIAG_ results, 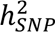 slightly decreased. We believe this is due to heterogeneous measurement error and bias in the ADHD_SYMP_ phenotyping (Wang et al., 2023). The same can be observed when looking at the differences in 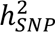 between ADHD_DIAG_ from 2019 (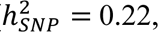 including 23andMe), which was strict in its definition of ADHD cases, and ADHD_DIAG_ from 2023 (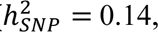 including 23andMe), which was slightly more lenient in its definition of ADHD cases.

MAGMA analyses identified 64 potential ADHD risk genes. These genes were significantly enriched in genes previously identified in GWASs of cognitive phenotypes and risk-taking behaviors. The total GWAS signal was significantly differentially expressed in several brain specific tissue types, general brain tissue types and the pituitary gland, as well as in late infancy Brainspan brain samples. FUMA gene-mapping mapped significant loci to 204 genes. Again, genes were enriched in gene-sets reported by previous GWASs on cognitive behavior, risk-seeking behavior, and brain development. FUMA enrichment analyses further revealed 29 transcription factor targets that may be of interest for ADHD.

To get a better understanding of the causal pathways from SNPs to ADHD, we ran FLAMES (Schipper et al., 2023) to identify likely effector genes. FLAMES identified 22 potential effector genes, of which 14 overlapped with the MAGMA genes, 12 were previously reported by Demontis et al. (2023) and 8 were not previously linked to ADHD or not reported on GWAS Catalog. The 22 genes were significantly overrepresented in gene-sets differentially expressed in the frontal cortex, enriched in 4 GO biological processes related to neural and physical development, 52 transcription factor targets, 13 microRNA targets, 8 different cell type signatures, 4 synapse cellular components, and 3 synaptic biological processes. In Demontis et al. (2023), the set of potential ADHD risk genes was significantly enriched among genes upregulated during early embryonic brain development, this result was not replicated in our study. A common theme is that implicated genes are enriched in processes that are involved in neural development and functioning. The results provide several new avenues to investigate, which could prove useful in gaining more insights into the etiology of ADHD. The results may provide more potentially useful information for the 22 potential effector genes compared to the 204 genes identified by FUMA positional mapping, eQTL mapping, and chromatin interaction mapping. We think this difference results from the difference in strategies employed by both methods. FUMA maps every gene for which some functional link is known to exist, whereas FLAMES weighs all these measurements and only prioritizes genes if they are clearly more likely causal genes than the other genes in the locus. Our findings suggests that FLAMES can help identify functional pathways that may remain hidden with other approaches due to a reduction of noise from non-causal genes in the set of prioritized genes, which decreases the power to detect enrichment in functional gene-sets.

Next, we examined genetic correlations between ADHD_overall_ and various external phenotypes. Except for anorexia nervosa, ADHD_overall_ showed significant genetic correlations with all examined psychopathological traits. Most striking was the genetic correlation of 1.13 with childhood aggressive behavior, and 0.97 with antisocial behavior. Previous studies reported moderate-to-strong phenotypic correlations across sex-, rater-, age-, and instrument-specific assessment between aggressive behavior and attention problems and hyperactivity (Bartels et al., 2018). We found a negative genetic correlation between ADHD and alchohol intake frequency, and a positive correlation between ADHD and number of drinks per week. Marees et al. (2020) studied similar correlations between alcohol consumption and mental health traits, and found evidence that suggests the different genetic correlations are a result of SES effects. We found a moderate genetic correlation with smoking behaviors, but a small correlation with cannabis use and discrepant findings for alcohol consumption. This was surprising, given the phenotypic associations that have been reported in previous studies (Lee et al., 2011). We found negative correlations with several cognitive traits, such as (childhood) IQ, verbal-numerical reasoning, and educational attainment. Similar to a GWAS of childhood aggression (Ip et al., 2021), genetic correlations with several hormone levels were around zero. Finally, we found a small negative correlation with birthweight, but a weak positive correlation with childhood obesity, and adult BMI. These genetic correlations suggest wide pleiotropic effects of variants involved with ADHD. This is illustrative of the polygenetic nature of most behavioral, cognitive, and (mental) health traits. It also indicates genetic factors play a role in the comorbidity of psychopathological disorders.

Combining data collected using different instruments and by different raters helps to increase the sample size, and with that the statistical power of our analyses. This is illustrated by the apparent increase in power to detect genetic variants associated with ADHD in the combination ADHD_SYMP_ and ADHD_DIAG_, compared to the combination ADHD_DIAG_ and EAGLE ADHD GWAS (Demontis et al., 2019; Middeldorp et al., 2016). However, this approach of including different raters and instruments also has some downsides. Correlations between raters and instruments are not always large, meaning that we introduce heterogeneous measurement error and bias in our phenotyping, which could suppress SNP-heritability. More homogeneous measurement may offer an avenue to higher powered GWAS. In light of increased collaborations between cohorts to expand sample sizes for GWAS, more efforts to standardize the assessment of traits could help further increase the power and specificity of GWAS results. Moreover, downstream analyses, for example using polygenic scores computed with the summary statistics of the current study, will favor cohorts using the same instruments and raters that dominated the discovery sample, in our case the Achenbach System of Empirically Based Assessment (Achenbach et al., 2017).

Assessments of ADHD in individuals from non-European ancestry were rare in each of the included cohorts. Because of the low number of assessments, we were forced to exclude non-European individuals from our analyses. We know that results from European ancestry GWASs often also significantly predict differences in non-European ethnic groups, but often effect sizes are diluted towards zero. Regrettably, this means that knowledge generated by these types of studies risks benefit individuals of European ancestry more than from diverse backgrounds. To better understand the etiology of ADHD across individuals and backgrounds, it is important to continue ongoing efforts to increase the inclusivity of GWAS samples.

In conclusion, the current study adds novel insight into the genetic etiology of ADHD. By meta-analyzing GWAS results from symptom counts of ADHD in children with a diagnosis of ADHD, we identified novel genome-wide significant loci and genes. The number of genome-wide (significant) genetic variants that are implicated in ADHD provides further insight into the polygenic etiology of ADHD. The 22 potential effector genes that were identified by FLAMES gives insight in several biological processes that may play a causal role in the etiology of ADHD, and provides avenues for further research. The genetic correlations with other phenotypes further indicate the wide pleiotropic effects of genetic variants and the role that genetic variants play in the co-occurrence with (mental) health traits.

## Methods

### Sample & cohorts

Childhood cohorts that collaborate within the ACTION (**A**ggression in **C**hildren: unraveling gene-environment interplay to inform **T**reatment and **I**nterventi**ON** strategies) consortium (Bartels et al., 2018; Boomsma, 2015; Ip et al., 2021) and EAGLE consortium (Middeldorp et al., 2019) took part in the meta-analysis of ADHD symptom counts (ADHD_SYMP_). Cohorts assessed ADHD Symptoms in children and adolescents aged 1.5 to 18 years and also included adult retrospective assessments. Each cohort received a standard operation protocol (see: https://www.action-euproject.eu/content/data-protocols). Cohorts could contribute one or several univariate genome-wide association studies (GWAS). A separate analysis was performed for every unique combination of rater, instrument, and age, with a minimum of 450 observations per GWAS (so that each GWAS included a maximum of one measure for each individual). Extended information on the cohorts can be found in Supplementary Table 1 and the Supplementary Text. Assessments of individuals of non-European ancestry were limited, and analyses were restricted to individuals of European ancestry. In total, 28 cohorts contributed 154 GWASs, resulting in a total of 288,887 observations from 72,483 unique individuals (Supplementary Table 2).

### Measurement of ADHD symptoms

ADHD symptoms in children and adolescents were rated by parents, teachers, and the individuals themselves. Additionally, retrospective assessments of (pre-) adolescent ADHD symptoms from self-report or maternal report were included in the GWAMA. To maximize sample size, we included measurement of ADHD symptoms across multiple instruments. In total, 22 ADHD symptom assessment instruments were included in the meta-analysis (Supplementary Table 3). The most commonly employed instruments were the Achenbach System of Empirically Based Assessment (Achenbach et al., 2017) and the Strengths and Difficulties Questionnaire (Goodman, 2001).

### Genotyping and quality control

Genotyping was performed within each cohort using common genotyping arrays (see Supplementary Table 4), followed by cohort-specific quality control based on individual- and variant-based call rate, Hardy-Weinberg equilibrium, excessive heterozygosity rates, and minor allele frequency (see Supplementary Table 5). Cohorts removed samples with non-European ancestry. 78.6% of the cohorts imputed their genotypes to 1000 Genomes (1000G) phase 3 version 5, while the remaining cohorts used 1000G phase 1 version 3 as the reference set for the imputation (Supplementary Table 6). All genotypes were mapped to build 37 of the Human Genome Reference Consortium assembly (GRCh37).

### GWA model

Each cohort performed a univariate GWAS where ADHD symptoms were regressed on the SNP genotype, with age, sex, and first five ancestry-based principal components as fixed effects, and, if necessary, cohort-specific covariates (Supplementary Table 7). To correct for dependency between observations within univariate analyses, cohorts with related individuals applied a mixed linear model (Tucker et al., 2015) or a sandwich correction of the standard errors (Minică et al., 2015).

GWASs were stratified by (1) rater, (2) instrument, and (3) age, so that observations within a univariate GWAS were independent, with a minimum stratum sample size of 450 observations. In total, summary statistics for 154 univariate GWAS were uploaded. Descriptive statistics for each uploaded GWAS are shown in Supplementary Table 8. Each cohort also supplied information on the degree of sample overlap and phenotypic correlation between their univariate analyses. These statistics allowed us to account for dependency between observations within cohorts.

### Pre-GWAMA QC

Summary statistics from each GWAS went through quality control (QC) using the EasyQC software package (Winkler et al., 2014). SNPs with a genotyping rate below 95% were removed. We applied variable QC filters on MAF and HWE *p*-value tailored to the sample size in the specific GWAS. Respective cutoffs of INFO> 0.6 and INFO> 0.7 were applied to SNPs that were imputed using MACH and IMPUTE (Liu et al., 2015). Reported allele frequencies were compared to the allele frequency in an imputation-matched reference population and variants with an absolute difference in allele frequency larger than 0.2 were removed. Supplementary Table 9 reports the number of SNPs before and after QC.

### Meta-analysis of ADHD symptoms

The meta-analysis method is equal to the method described by Ip and colleagues (Ip et al., 2021). Due to sample overlap between the univariate GWASs, covariance between GWAS test statistics are a function of sample overlap and a truly shared genetic signal (Bulik-Sullivan, Loh, et al., 2015). To correct for sample overlap during the meta-analysis, we applied a modified version of the multivariate meta-analysis approach developed by Baselmans and colleagues (2019), where we calculated the expected cross-trait-intercept (Bulik-Sullivan, Loh, et al., 2015) based on the observed sample overlap and phenotypic covariance, as reported by the cohorts. Finally, because the sum of the number of observations (N_obs_) was an overestimate of the effective sample size (N_eff_), we approximated the effective sample size as proposed by Ip and colleagues (2021) 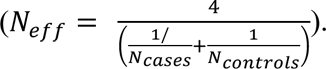 SNPs with MAF< 0.01, N_eff_< 15,000, or were observed in only one cohort were removed from further analyses. SNP-heritability (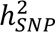) was estimated by genomic Structural Equation Modeling in R (Grotzinger et al., 2019).

### Meta-analysis with case-control ADHD GWAS

In the next step, we meta-analyzed our ADHD_SYMP_ GWAMA with a GWAS of ADHD diagnosis (Demontis et al., 2023). In ADHD_DIAG_, cases are defined as clinically diagnosed with ADHD or prescribed medication specific for ADHD. ADHD_DIAG_ included data from the Lundbeck Foundation Initiative for Integrative Psychiatric Research (iPSYCH), the Psychiatric Genomics Consortium (PGC), and deCode. Data were obtained for adults and children, resulting in a total of 38,691 cases and 186,843 controls.

For the meta-analysis, we first adjusted the test statistics and sample sizes for ADHD_SYMP_ and ADHD_DIAG_ as proposed by (Demontis et al., 2019). The lifetime population prevalence of ADHD was assumed to be 5% (Faraone et al., 2015). SNP-heritability for ADHD_SYMP_, and ADHD_DIAG_, and *r*_g_ and cross-trait intercepts (CTI) between ADHD_SYMP_ and ADHD_DIAG_ were estimated by genomicSEM in R (Grotzinger et al., 2019). We meta-analyzed the results from ADHD_SYMP_ and ADHD_DIAG_ based on the approach outlined in Baselmans and colleagues (Baselmans et al., 2019). We specified the effective sample sizes for ADHD_DIAG_ as suggested by Ip and colleagues (2021): 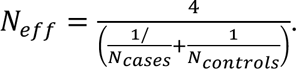 SNP-heritability was estimated using LDSC in genomicSEM (Grotzinger et al., 2019). We assumed no sample overlap between ADHD_SYMP_ and ADHD_DIAG_.

### Follow-up analyses

#### Fine mapping & gene-based tests

We used FUMA (Watanabe et al., 2017) to identify independent genome-wide significant loci and associated genes. LD blocks of independent significant SNPs within 250kb were merged into a single genomic locus. Protein-coding genes were mapped if they were located within a maximum distance of 10 kb of an independent significant SNP, or if a credible variant was annotated to the gene based on eQTL data or chromatin interaction data from human brain (see Supplemental Text for the included datasets). These are the same settings used by Demontis et al. (Demontis et al., 2023). Gene-based tests were run in MAGMA (de Leeuw et al., 2015). MAGMA gene-based tests combine *P*-values from multiple SNPs inside a gene to obtain a test statistic for each gene (Z_gene_), while accounting for incomplete linkage disequilibrium between SNPs. To this end, a list of 18,296 genes and their start- and end-positions, and pre-formatted genotypes, based on 1000G phase 3, were obtained from the MAGMA website (see Supplements). We applied a Bonferroni correction for multiple testing at α= 0.05 / 18,296 = 2.733 x 10^-6^.

It remains difficult to identify which genes are causally involved in ADHD. Fine-mapped Locus Assessment Model of Effector geneS (FLAMES) was recently developed with the goal of predicting the most likely effector genes from GWAS results. FLAMES is a novel framework that combines SNP-to-gene evidence and convergence-based evidence, outputting a single score per gene from fine-mapped GWAS loci. We performed statistical fine-mapping using FINEMAP version 1.4.1 (Benner et al., 2016), and an LD reference panel of 100,000 unrelated UK biobank participants of European descent. Given that the GWAMA contains cohorts that do not belong to the UK biobank we restricted the maximum number of causal variants in a locus modelled by FINEMAP to 1, to avoid overfitting. We ran FLAMES (version 1.0.0) by inputting pathway naïve PoPS scores (Weeks et al., 2023) for our GWAMA, the FUMA defined loci and corresponding fine-mapped credible sets, resulting in a single FLAMES score per gene. Genes with FLAMES scores above 0.05 were interpreted as potential effector genes, as suggested by the FLAMES authors. For more information on FLAMES, and the included functional annotations, see Schipper et al. (2023). Functional annotation and enrichment analysis was done for a set of genes with FLAMES scores above 0.05.

#### Enrichment & pathway analyses

We performed MAGMA gene-set analyses using the full ADHD_overall_ results. Gene property analysis was performed to test relationships between tissue specific gene expression profiles (See Supplementary Notes for an overview) and ADHD-gene associations. Next, genes mapped by FUMA, and the set of potential effector genes identified with FLAMES were used in gene-set enrichment analyses. We ran hypergeometric tests using FUMA genes2func to assess if genes of interest are overrepresented in any of the pre-defined gene sets (See Supplemental Notes for all included gene-sets). We used SynGO (Koopmans et al., 2019) v1.2 (“20231201”) to test for enrichment in genes encoding for proteins involved in synaptic cellular components and biological pathways. The brain expressed background set was used, containing 18,035 unique genes.

#### Genetic correlations

We computed genetic correlations between ADHD_overall_ and 49 preselected traits, including cognition and externalizing behaviors, psychopathologies, anthropometric measures, metabolic, and health outcomes (see Supplementary Table 10). Phenotypes were selected based on established hypotheses or were at least nominally significantly (*P*<0.05) genetically correlated with ADHD_DIAG2019_ (Demontis et al., 2019). Following Bulik-Sullivan et al. (Bulik-Sullivan, Finucane, et al., 2015) we restricted genetic correlations to external phenotypes for which the *Z*-scores of the LDSC-based 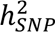 are ≥ 4.

## Data Availability

All data produced are available online at <link to follow>

## Acknowledgements

We very warmly thank all participants, their parents, and teachers for making this study possible. The project was supported by the *Aggression in Children: Unraveling gene-environment interplay to inform Treatment and InterventiON strategies* project (ACTION). ACTION received funding from the European Union Seventh Framework Program (FP7/2007-2013) under grant agreement no 602768. Cohort-specific acknowledgements and funding information are included in the Supplementary text.

## Ethics & Inclusion Statement

This study is the result of a large collaborative effort among multiple clinical and population-based cohorts. Researchers and principal investigators (PIs) representing the individual cohorts were involved in the design and execution of the study. Cohort specific GWAS analyses were performed locally, by local researchers. Local researchers and PIs were included as co-authors in consultation with the PIs of each included cohort. Data collections for the cohorts were approved by local ethics committees. Study approval was obtained from the Central Ethics Committee on Research Involving Human Subjects of the VU University Medical Center, Amsterdam (NTR 25th of May 2007 and ACTION 2013/41 and 2014.252), an Institutional Review Board certified by the U.S. Office of Human Research Protections (IRB number IRB00002991 under Federal-wide Assurance- FWA00017598; IRB/institute codes).

## URLs

GWAS Catalog: https://www.ebi.ac.uk/gwas/

FUMA: https://fuma.ctglab.nl

SynGO: https://www.syngoportal.org

Summary statistics: TBA

## Code & Summary Statistics

Analysis plan: https://www.action-euproject.eu/sites/default/files/Action%20AGG%20AP%20SOP.pdf

Code: -link to follow-

Summary Statistics: -link to follow-

