## Supplemental Figures for "Meta-analysis of Genome wide Association Studies on Childhood ADHD Symptoms and Diagnosis Reveals 17 Novel Loci and 22 Potential Effector Genes"

#### MAGMA

**Figure 1.** MAGMA genes Manhattan plot.

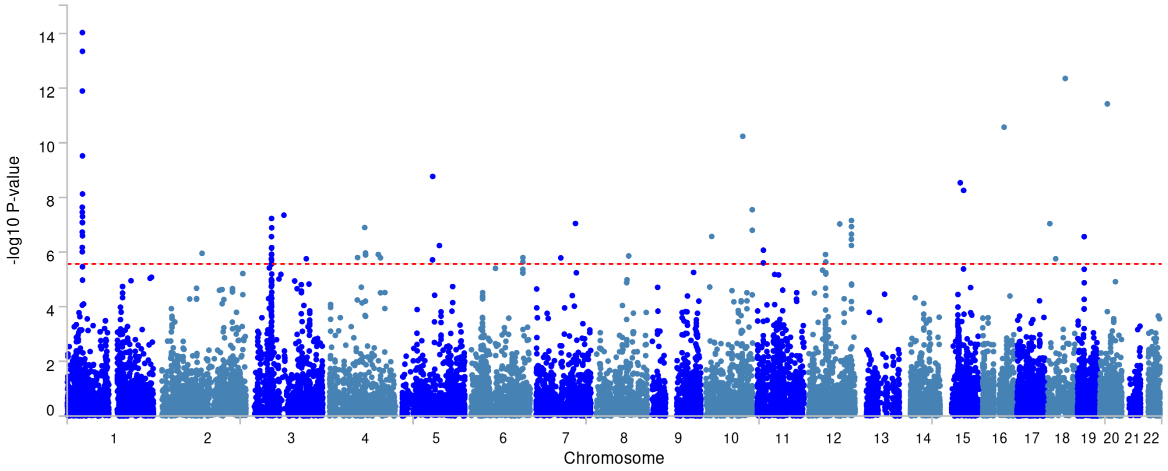


*Note:* Red line reflects genome-wide significance threshold (*P* = 0.05/18296= 2.73e-6).

**Figure 2 (a,b,c,d).** Results from MAGMA gene property tissue expression analyses

**a)** GTEx v8 specific tissue types

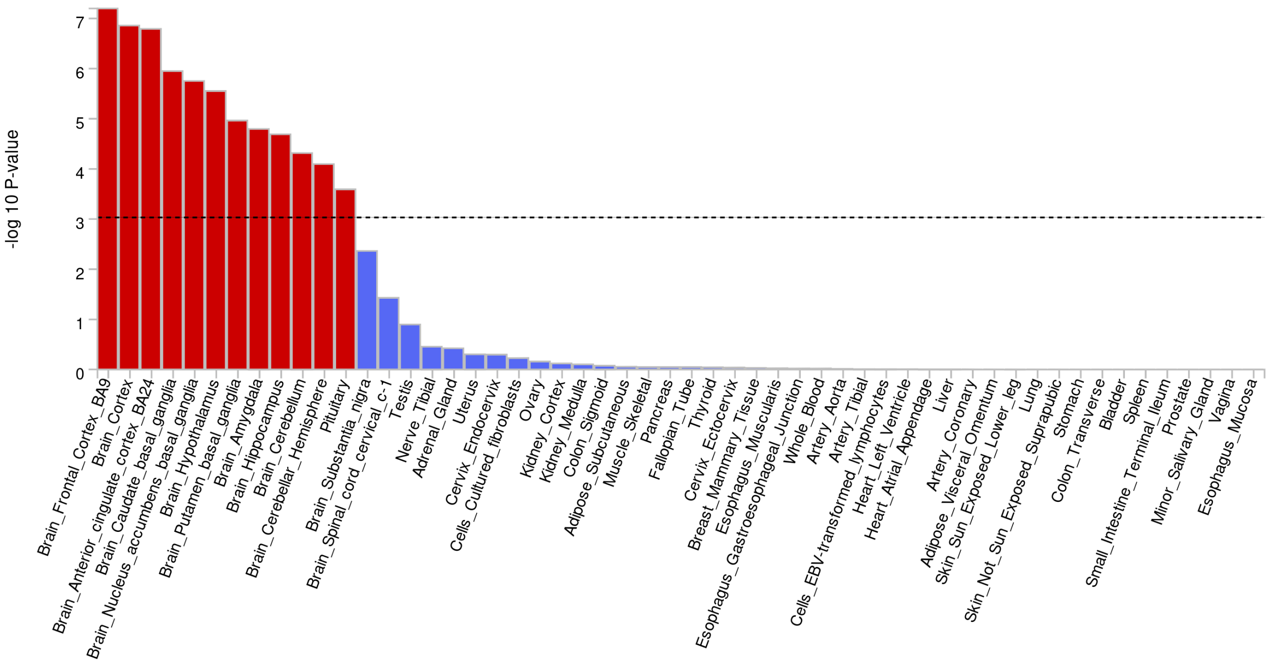


**b)** GTEx v8 general tissue types

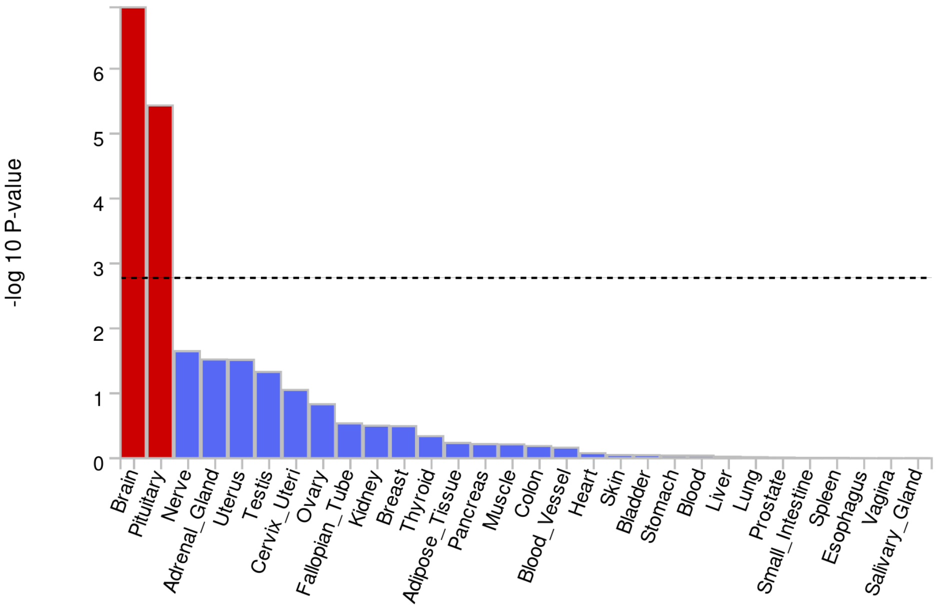

**c)** Brainspan 11 developmental stages of brain samples

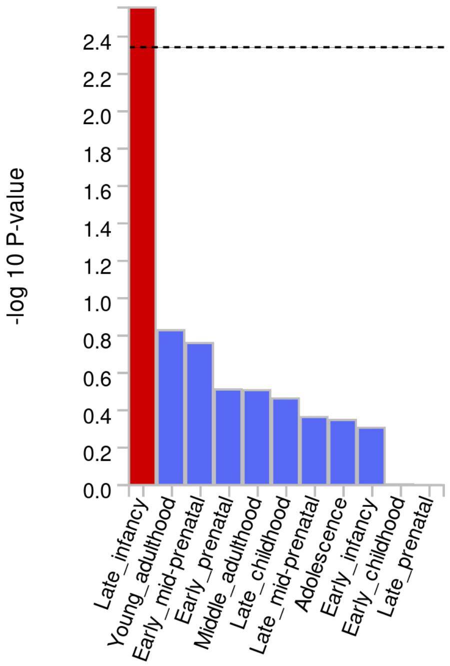


**d)** Brainspan 29 ages of brain samples

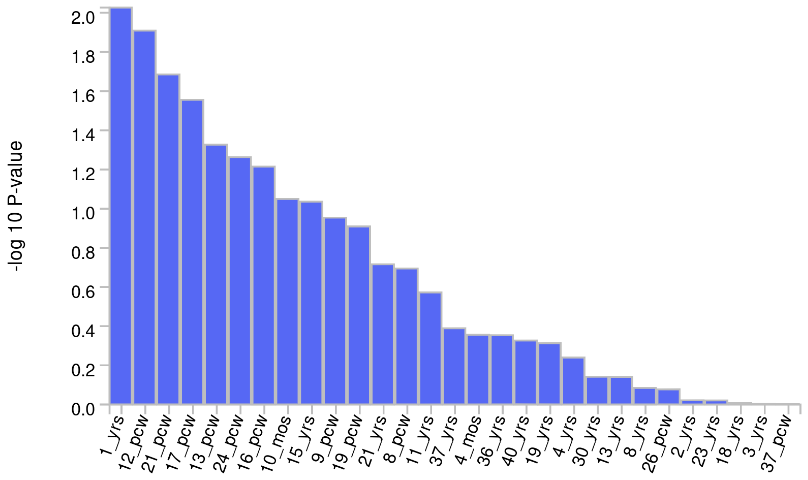


*Note:* To identify tissue specificity of ADHD, FUMA performs MAGMA gene-property analyses to test relationships between tissue specific gene expression profiles and ADHD-gene associations. Results are one-sided -log10 P-values. Expression profiles include GTEx v8 specific and general tissue types, Brainspan 11 developmental stages and 29 ages of brain samples. The test compares gene-expression per tissue type to the average expression across all categories. Red bars indicate significant results (Bonferroni corrected *P*-value $\leq 0.05$).

**FUMA mapped genes**

**Figure 3.** Results from hypergeometric tests of overrepresentation of FUMA mapped genes in gene sets from GWASs listed on GWAS Catalog

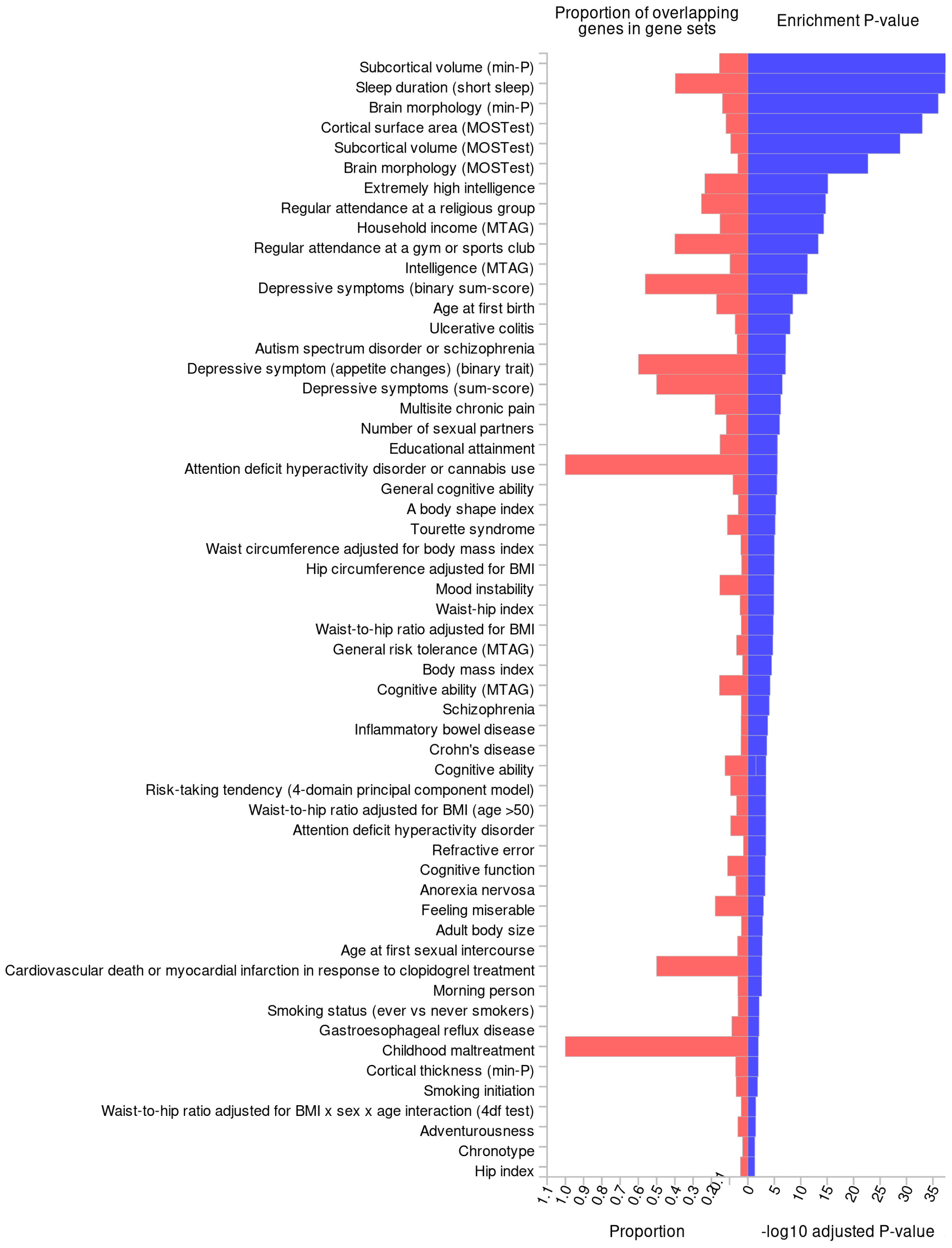


*Note:* The GWAS catalog data was downloaded with gene symbols and then converted to entrez ID using biomaRt. If a single gene symbol matched multiple entrez IDs, then all matching entrez IDs were included in the gene set.

**Figure 4 (a,b,c,d).** FUMA tissue specificity tests

**a)** GTEx v8 specific tissue types

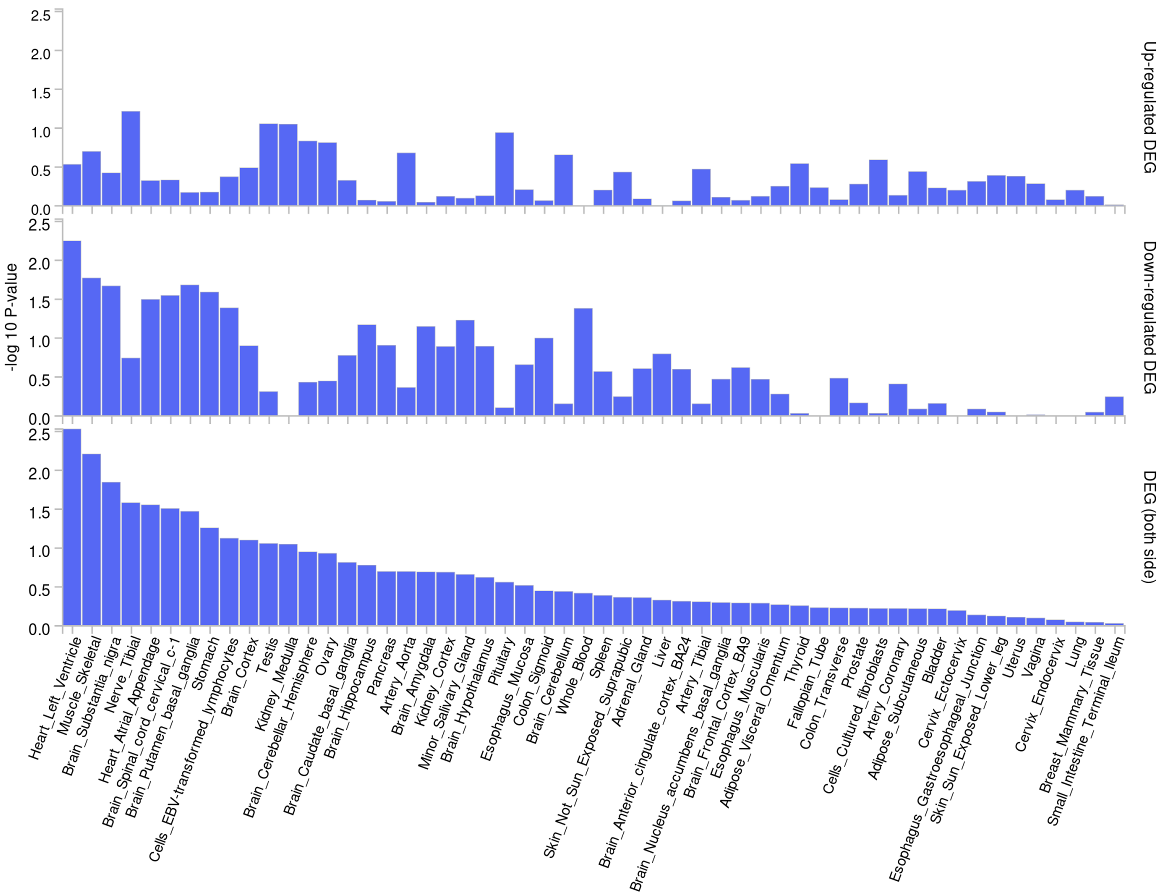

**b)** GTEx v8 general tissue types

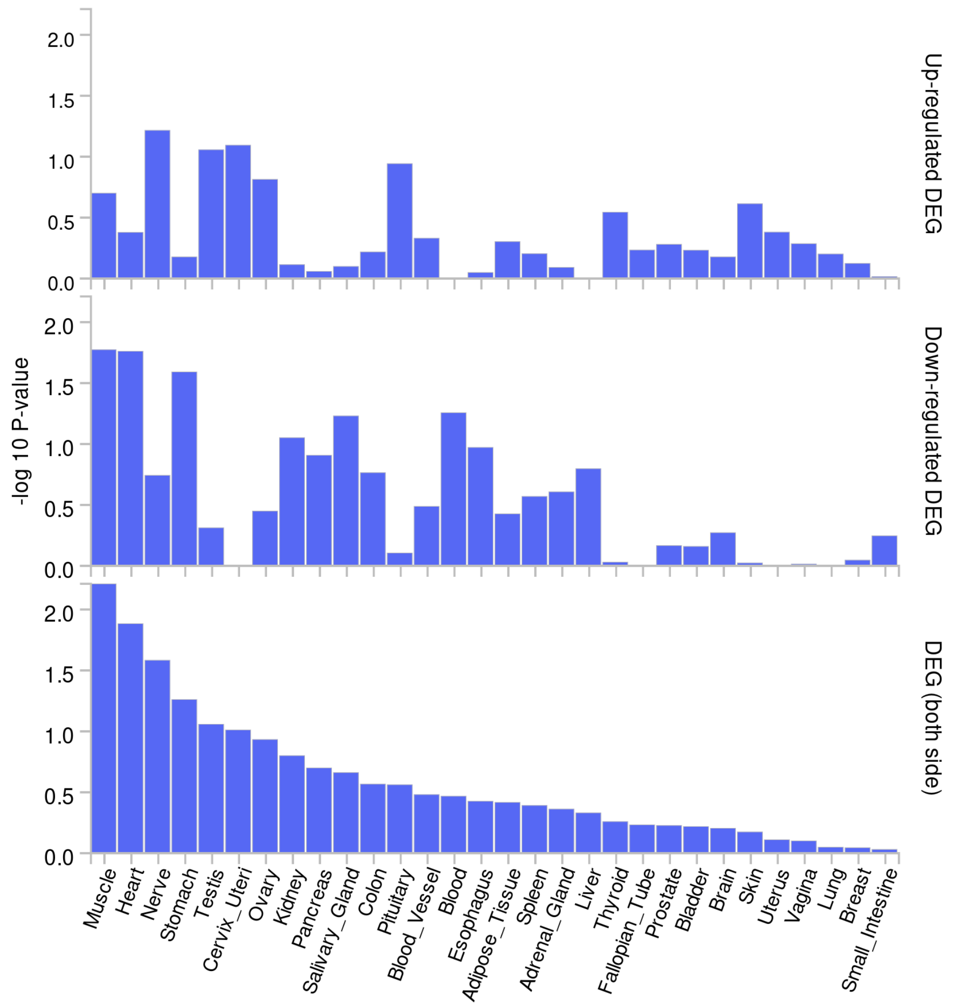

**c)** Brainspan 11 developmental stages of brain samples

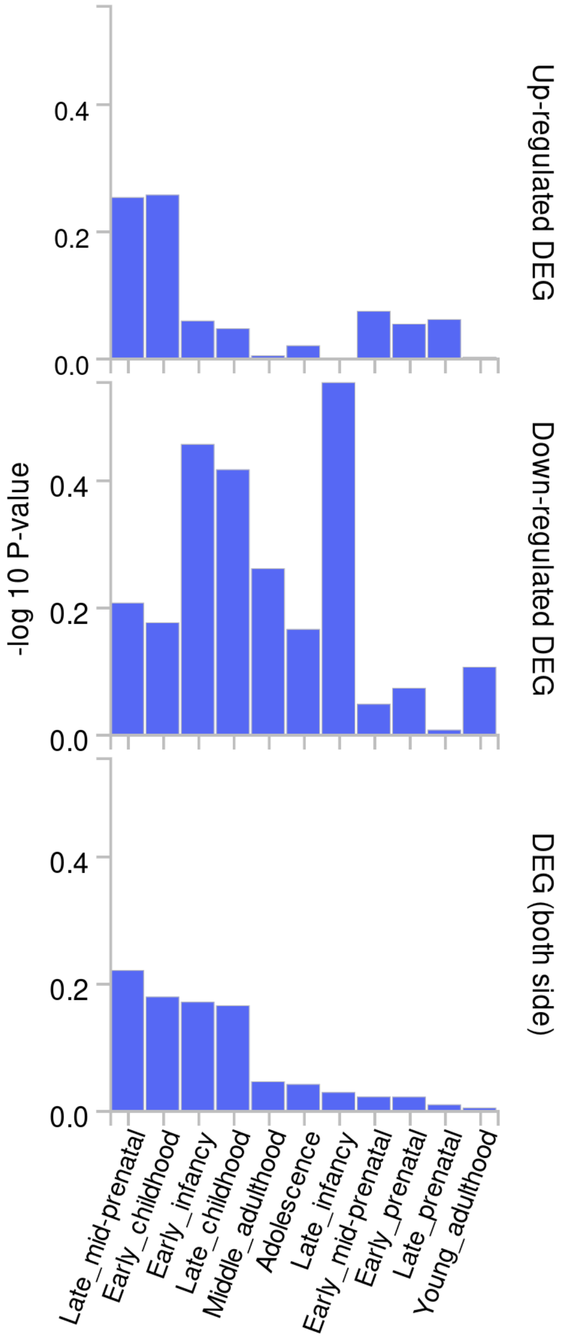

**d)** Brainspan 29 ages of brain samples

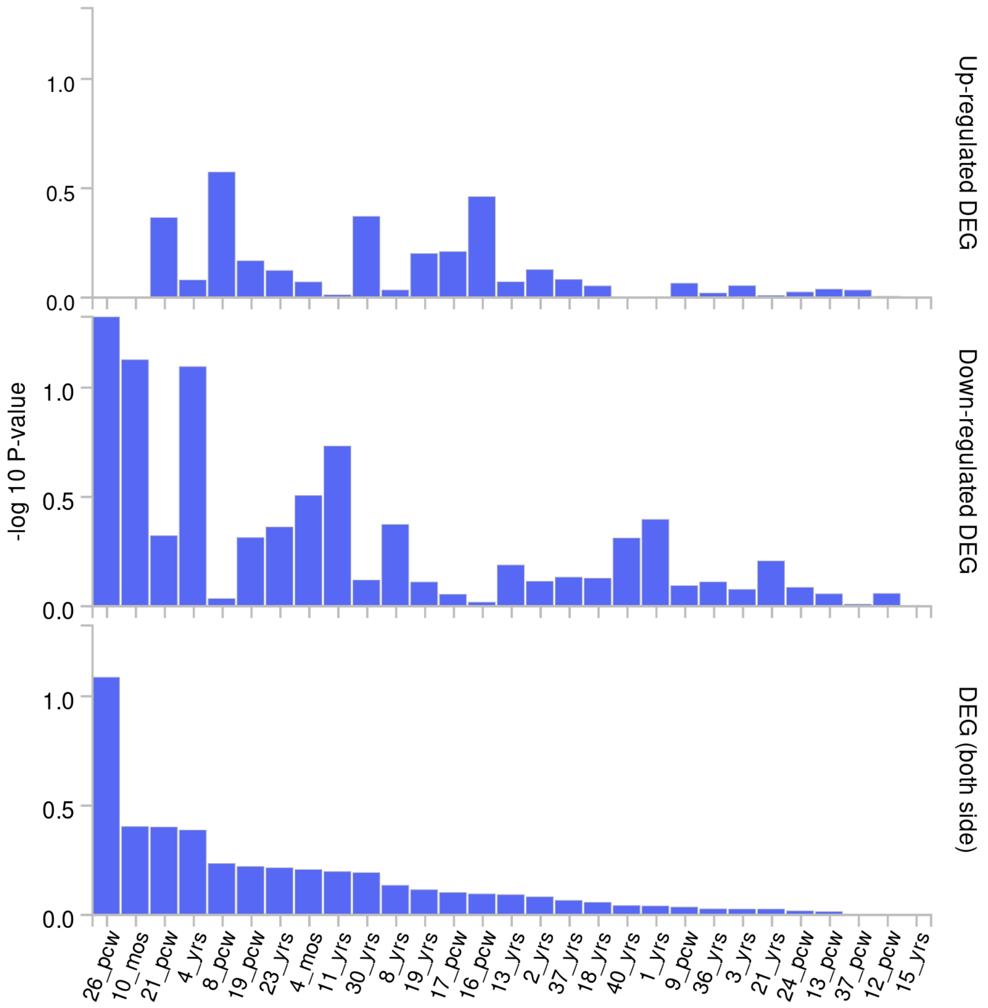


*Note:* Tissue specificity was tested using the differentially expressed genes defined for each label of each expression data set. Input genes were tested against each of the DEG sets using the hypergeometric test. The background genes were genes that had average expression values > 1 in at least one of the labels and existed in the selected protein-coding genes. Significant enrichment at Bonferroni corrected P-value ≤ 0.05 are colored in red. Bonferroni correction is performed for each up-regulated, down-regulated and both-sided DEG sets separately. Tissue specificity DEG sets included GTEx v8 specific and general tissue types, Brainspan 11 developmental stages and 29 ages of brain samples.

**Figure 5.** Results from hypergeometric tests of overrepresentation of FUMA mapped genes in gene sets that code for transcription factor targets.

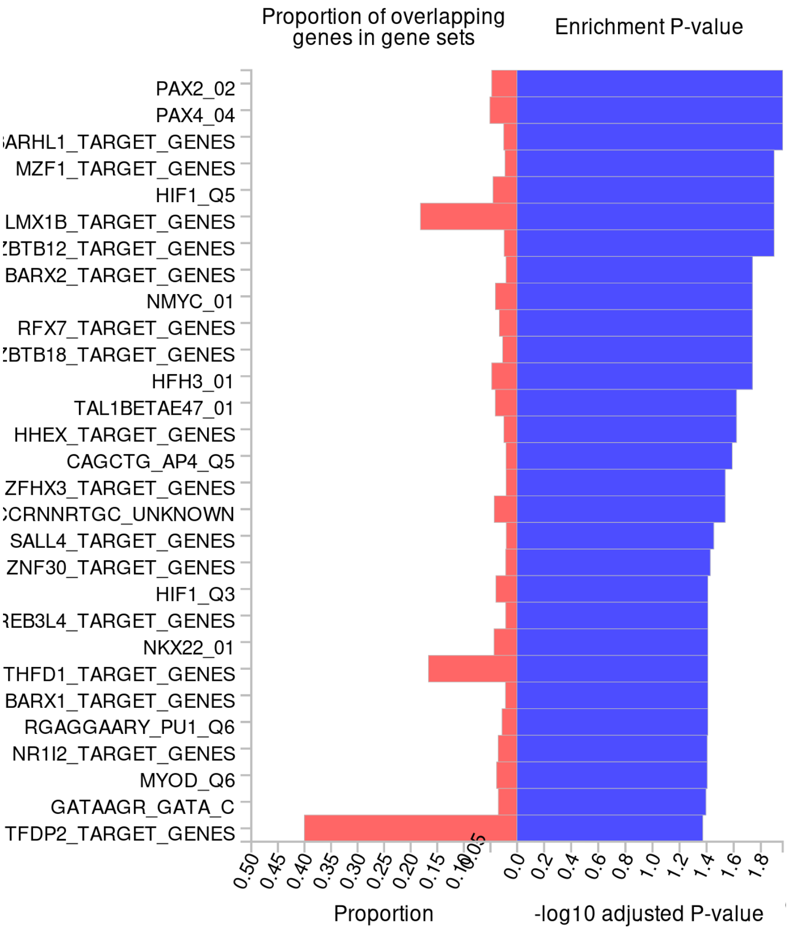


*Note:* Results (-log10 Bonferroni corrected *P*-values) from hypergeometric tests of overrepresentation of FUMA mapped ADHD genes in gene-sets that code for transcription factor targets. Only gene-sets in which ADHD genes are overrepresented after Bonferroni correction are presented.

### FLAMES effector genes

**Figure 6.** Results from hypergeometric tests of overrepresentation of 22 FLAMES effector genes in gene sets from GWASs listed on GWAS Catalog

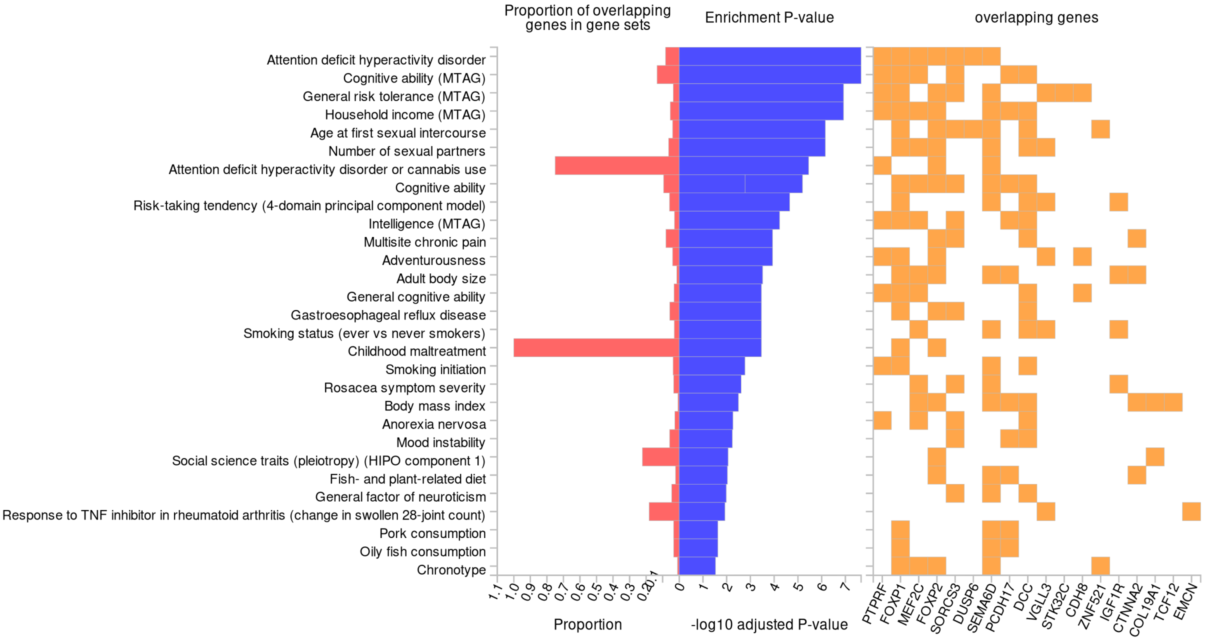


*Note:* The GWAS catalog data was downloaded with gene symbols and then converted to entrez ID using biomaRt. If a single gene symbol matched multiple entrez IDs, then all matching entrez IDs were included in the gene set.

**Figure 7 (a,b,c,d)**. FUMA tissue expression analyses of 22 FLAMES effector genes (GTEx v8 specific and general tissue types, Brainspan 11 developmental stages and 29 ages of brain samples).

**a)** GTEx v8 specific tissue types

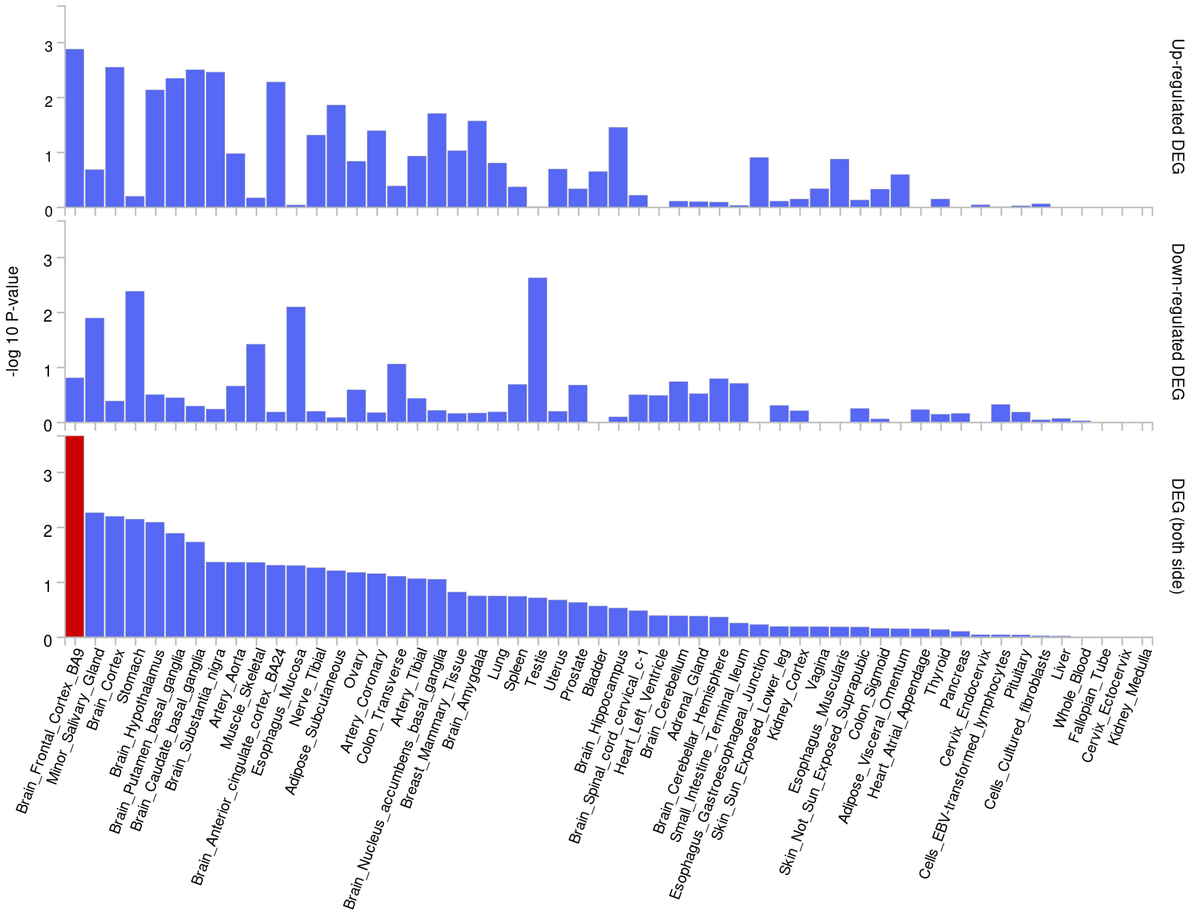

**b)** GTEx v8 general tissue types

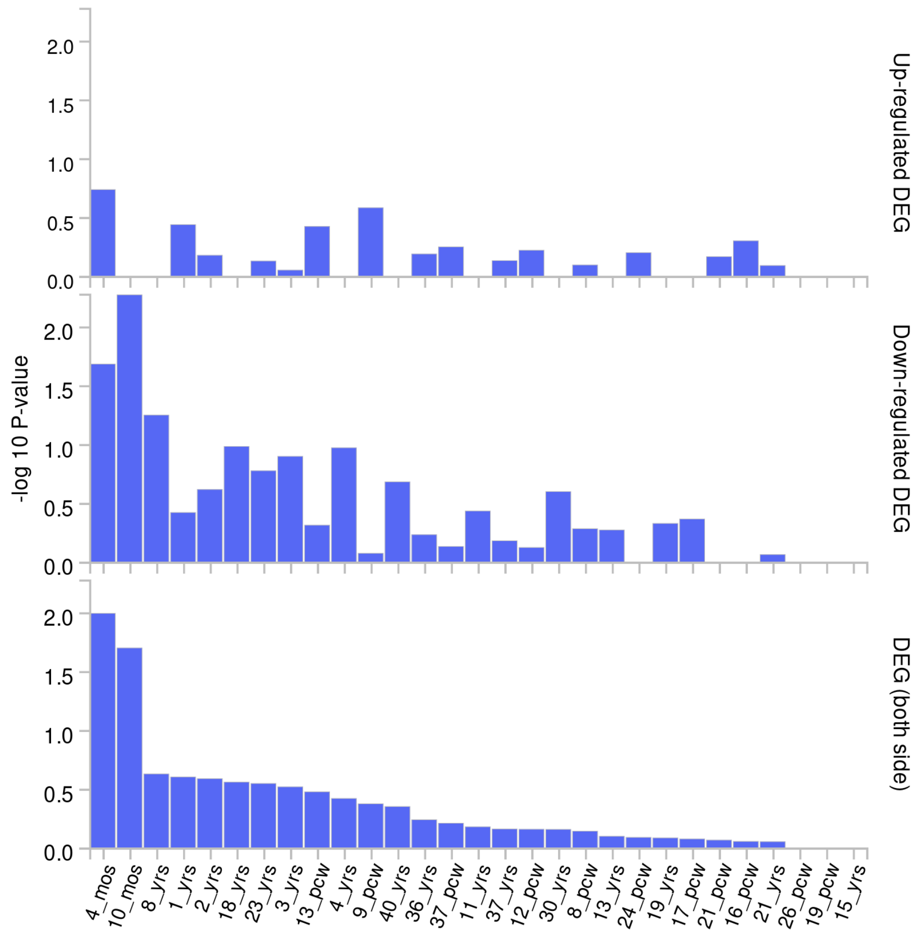

**c)** Brainspan 11 developmental stages of brain samples

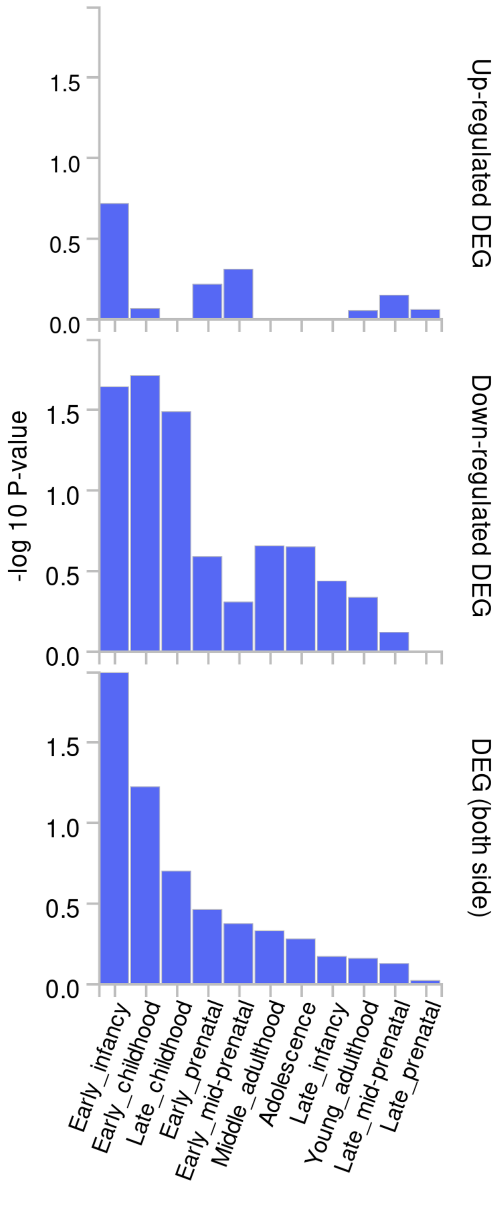

**d)** Brainspan 29 ages of brain samples

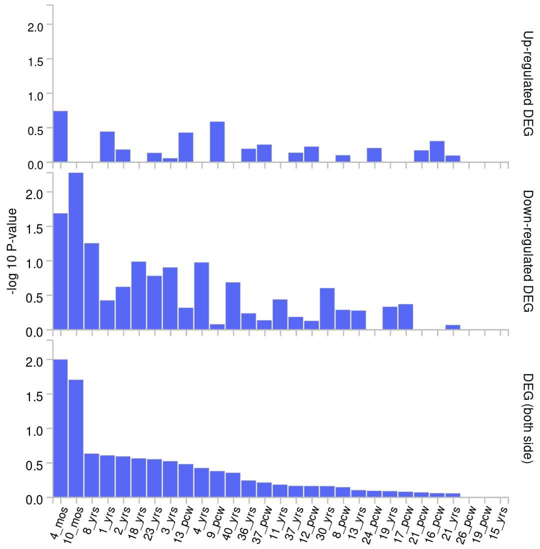


*Note:* Tissue specificity was tested using the differentially expressed genes defined for each label of each expression data set. Input genes were tested against each of the DEG sets using the hypergeometric test. The background genes were genes that had average expression values > 1 in at least one of the labels and existed in the selected protein-coding genes. Significant enrichment at Bonferroni corrected P-value ≤ 0.05 are colored in red. Bonferroni correction is performed for each up-regulated, down-regulated and both-sided DEG sets separately. Tissue specificity DEG sets included GTEx v8 specific and general tissue types, Brainspan 11 developmental stages and 29 ages of brain samples.

**Figure 8.** Results from hypergeometric tests of overrepresentation of FLAMES effector genes in gene sets that code for transcription factor targets.

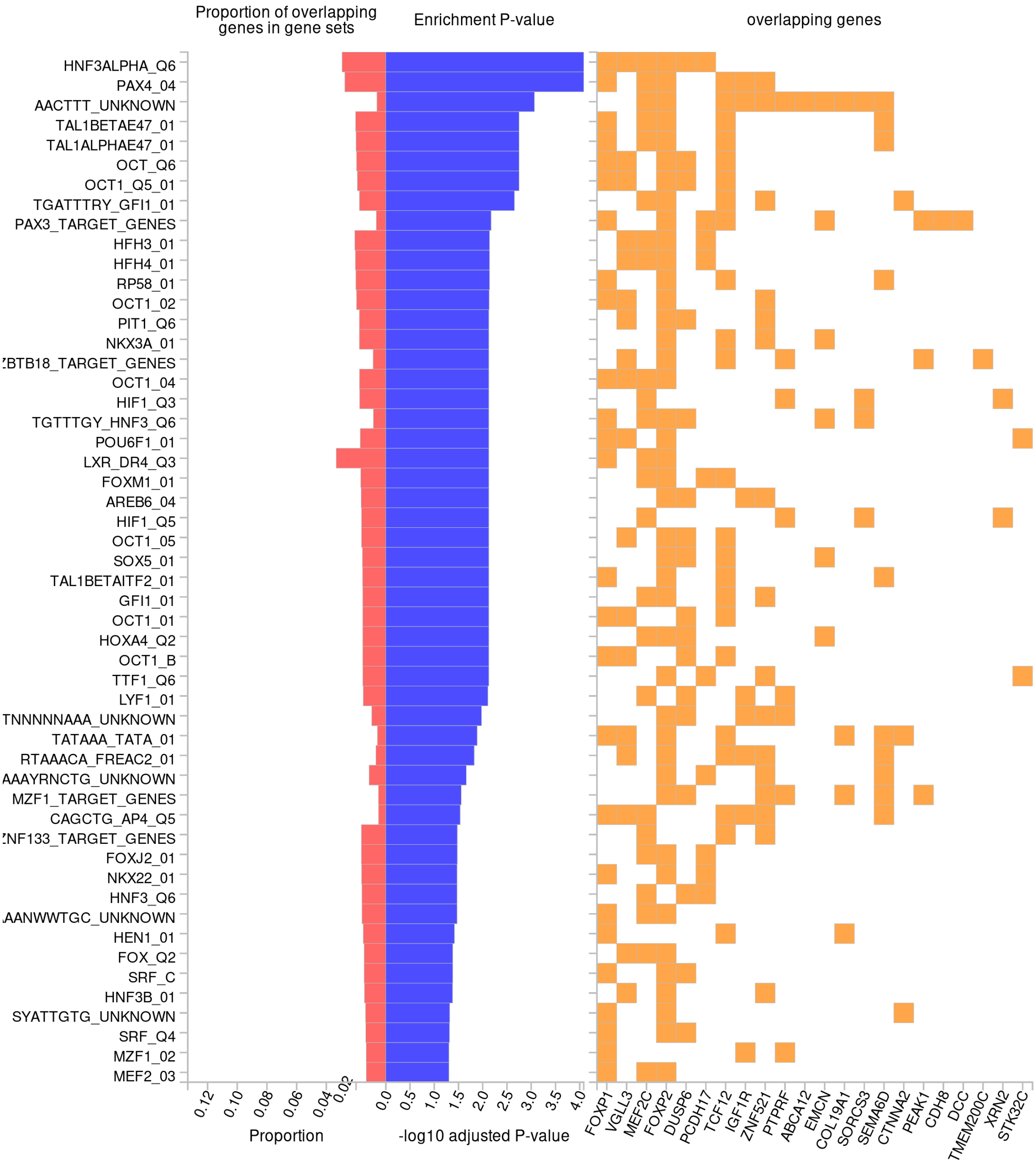


*Note:* Results (-log10 Bonferroni corrected *P*-values) from hypergeometric tests of overrepresentation of FLAMES ADHD effector genes in gene-sets that code for transcription factor targets. Only gene-sets in which ADHD genes are overrepresented after Bonferroni correction are presented.

**Figure 9.** Results from hypergeometric tests of overrepresentation of FLAMES effector genes in gene sets coding for GO Biological processes.

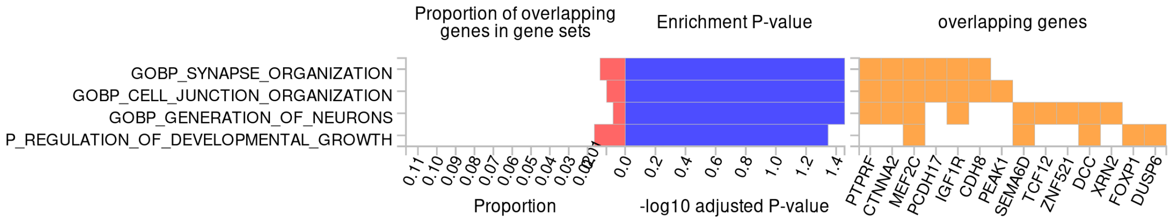


*Note:* Results (-log10 Bonferroni corrected *P*-values) from hypergeometric tests of overrepresentation of FLAMES ADHD effector genes in gene-sets that code for GO Biological processes. Only gene-sets in which ADHD genes are overrepresented after Bonferroni correction are presented.

**Figure 10.** Results from hypergeometric tests of overrepresentation of FLAMES effector genes in gene sets that are involved in canonical pathways.

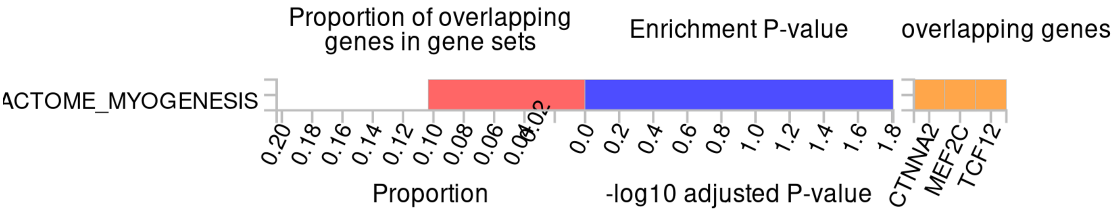


*Note:* Results (-log10 Bonferroni corrected *P*-values) from hypergeometric tests of overrepresentation of FLAMES ADHD effector genes in gene-sets that that are involved in canonical pathways. Only gene-sets in which ADHD genes are overrepresented after Bonferroni correction are presented.

**Figure 11.** Results from hypergeometric tests of overrepresentation of FLAMES effector genes in microRNA target gene-sets.

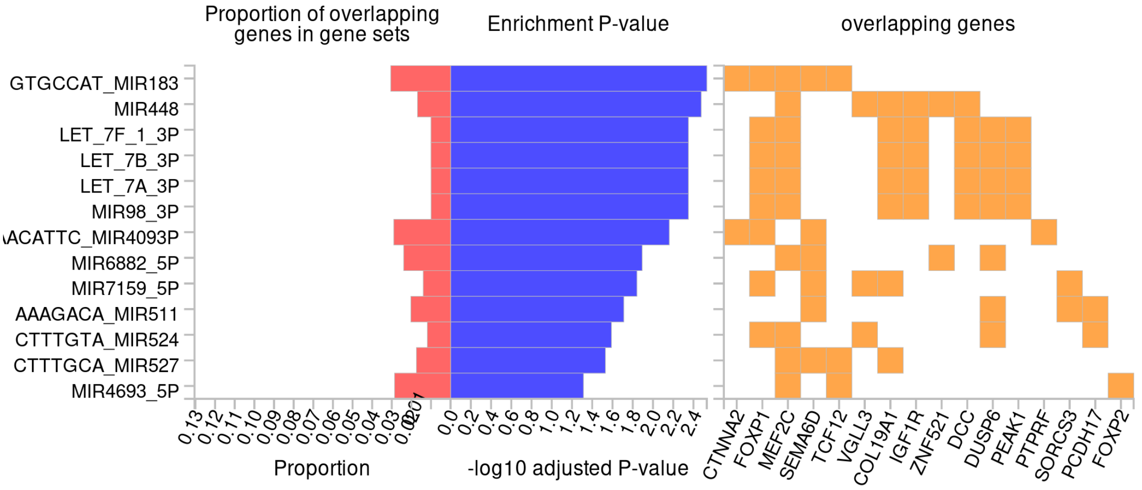


*Note:* Results (-log10 Bonferroni corrected *P*-values) from hypergeometric tests of overrepresentation of FLAMES ADHD effector genes in microRNA target gene-sets. Only gene-sets in which ADHD genes are overrepresented after Bonferroni correction are presented.

**Figure 12.** Results from hypergeometric tests of overrepresentation of FLAMES effector genes in cell-type signature gene-sets.

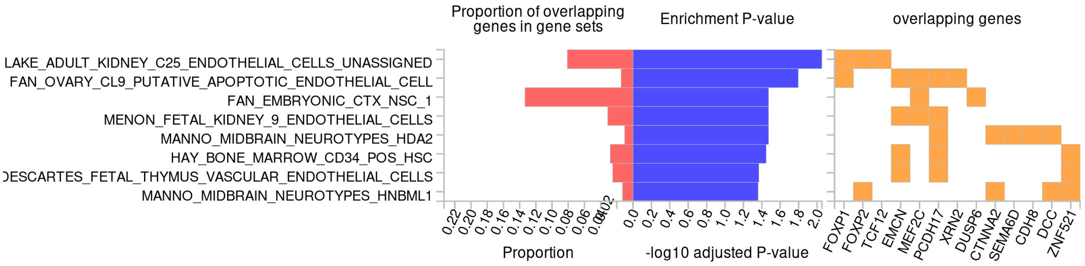


*Note:* Results (-log10 Bonferroni corrected *P*-values) from hypergeometric tests of overrepresentation of FLAMES ADHD effector genes in cell-type signature gene-sets. Only gene-sets in which ADHD genes are overrepresented after Bonferroni correction are presented.
